# A Gold Standard Dataset for Lineage Abundance Estimation from Wastewater

**DOI:** 10.1101/2024.02.15.24302811

**Authors:** Jannatul Ferdous Moon, Samuel Kunkleman, William Taylor, April Harris, Cynthia Gibas, Jessica Schlueter

**Affiliations:** SARS-COV-2; Wastewater; Deconvolution; Benchmark; Control Dataset; Deconvoluting tools; Relative abundance

## Abstract

During the SARS-CoV-2 pandemic, genome-based wastewater surveillance sequencing has been a powerful tool for public health to monitor circulating and emerging viral variants. As a medium, wastewater is very complex because of its mixed matrix nature, which makes the deconvolution of wastewater samples more difficult. Here we introduce a gold standard dataset constructed from synthetic viral control mixtures of known composition, spiked into a wastewater RNA matrix and sequenced on the Oxford Nanopore Technologies platform. We compare the performance of eight of the most commonly used deconvolution tools in identifying SARS-CoV-2 variants present in these mixtures. The software evaluated was primarily chosen for its relevance to the CDC wastewater surveillance reporting protocol, which until recently employed a pipeline that incorporates results from four deconvolution methods: Freyja, kallisto, Kraken2/Bracken, and LCS. We also tested Lollipop, a deconvolution method used by the Swiss SARS-CoV2 Sequencing Consortium, and three recently-published methods: lineagespot, Alcov, and VaQuERo. We found that the commonly used software Freyja outperformed the other CDC pipeline tools in correct identification of lineages present in the control mixtures, and that the newer method VaQuERo was similarly accurate, with minor differences in the ability of the two methods to avoid false negatives and suppress false positives. These results provide insight into the effect of the tiling primer scheme and wastewater RNA extract matrix on viral sequencing and data deconvolution outcomes.

**Highlights:**

- Generation of a gold standard dataset
- Comparative evaluation of relative abundance estimation software
- Evaluation of deconvolution methods used in CFSAN’s CWAP pipeline

## 1. Introduction

SARS-CoV-2 emerged in China in December of 2019 and led to the COVID-19 pandemic [1]. From a public health perspective, as SARS-CoV-2 has continued to mutate, tracking circulating and emerging variants of SARS-CoV-2 has been an essential part of the pandemic response [2]. Sequencing-based wastewater surveillance has become a sentinel for monitoring and identifying these new variants and tracking the shifts in variants across populations [3]. During outbreaks, COVID-19 test samples have been plentiful and provided an adequate basis for identifying the emergence and introduction of new variants. However, as governments have changed or ended their commitments to COVID-19 surveillance, PCR testing has been replaced by at-home testing and the few clinical samples available only offer a sampling of individuals who either choose to be tested or are hospitalized.

Wastewater-based epidemiology is not a new concept; it has been used since the early 20th century for monitoring other outbreak-causing pathogens [4][5]. When early reports in 2020 showed that SARS-CoV-2 was detectable in wastewater and was a leading indicator in advance of spikes in confirmed cases [6], this sparked widespread interest in wastewater-based epidemiology (WBE) as a tool for monitoring COVID-19 outbreaks. Wastewater surveillance was implemented on a large scale worldwide, in locations ranging in size from large metropolitan sewersheds [7][8] to individual college campus dormitories [9][10]. The relatively low implementation cost of genome-based wastewater surveillance makes it ideal for areas that lack resources for clinical sample-based sequencing surveillance [11] and a useful addition to any areas where clinical sequencing is limited. In response to the COVID-19 pandemic, the CDC launched the National Wastewater Surveillance System (NWSS) in September 2020 [12] and the U.S. Food & Drug Administration [13] set up a sequencing project with the collaboration of national and university labs to track and monitor the incidence of SARS-CoV-2. The Center for Food Safety and Applied Nutrition of the FDA (2013) made their in-house pipeline, C-WAP, available to collaborating labs that submit wastewater sequencing to the NWSS [14]. As clinical testing has decreased since 2022, this approach gives epidemiologists and public health officials a means to track the proportion of variants circulating in the community, and can potentially identify new and novel variants as they emerge [15]. Recently, the C-WAP pipeline has been archived and is not under active development or maintenance; its successor Aquascope can be used instead and relies primarily on Kraken2 and Freyja, both of which are evaluated in this manuscript [14].

The most widely used approach for sequencing SARS-CoV-2 viral RNA was developed by the ARTIC Network [16]. This method uses tiled amplicon-based sequencing [3] to cover the whole genome using 300-500 bp amplicons which are then pooled and sequenced. The full process of wastewater sequencing includes sample concentration, RNA extraction, and target amplification out of the total extracted RNA from each sample [17]. Along with the ARTIC Network’s primers, several other amplicon primer sets exist to sequence SARS-CoV-2 and generate full coverage consensus sequences [18][19][20]. A second approach employs target enrichment, amplifying only the spike protein regions where many of the identifying mutations occur [21]; the proceeding concentration and extraction steps are the same regardless of the amplification approach.

Assembly of amplified viral genomic RNA is achieved by realigning amplicon sequences to a reference genome. The bioinformatic analysis of SARS-CoV-2 sequencing from wastewater is significantly more complicated than it is for clinical samples. With a clinical sample, it is reasonable to assume that an individual is going to be infected with only one variant. The bioinformatics approach to a clinical sample is to align all the amplicons to the reference and then identify the variant calls using standard approaches like minimap and BCFtools [22][23].

Wastewater samples are composite collections from the population served by a single building, sewershed, or wastewater treatment facility. Therefore the expectation is that the sample itself is a representation of the variants circulating in that community, and the nature and complexity of the mixture may vary with the population size in the area collected. When wastewater samples are analyzed, rather than simply aligning the sequences collected to a reference, they must be deconvoluted and assigned to specific variants. There are several bioinformatic pipelines available for the analysis of wastewater sequencing data. As with other metagenomic samples, wastewater sequencing comes with a variety of challenges. The chemical environment of wastewater leads to viral RNA degradation and fragmentation [24]. Low viral concentration requiring an initial sample concentration step is the norm in wastewater samples, and the concentration method during processing can also impact the coverage and quality of sequence data [3]. Wastewater also contains both low-frequency and high-frequency variants simultaneously, which can make the detection and relative abundance estimation of the lineages more difficult [11], especially since low-abundance components may still be of interest as new variants enter a monitored area. The current bioinformatics tools can often identify the dominant lineage but fail to detect low-frequency or new variants correctly. For these reasons, deconvolution of SARS-CoV-2 variants from mixed viral samples still presents a challenge [3].

One of the most commonly used tools for wastewater variant sequencing is Freyja [3]. It uses a depth weighted least absolute deviation regression algorithm and reports the relative lineage abundances in mixed viral samples mapping to a common viral reference from a sequencing dataset [25]. Using .bam files, it first calculates the frequency of each mutation and its respective sequencing depth. Then, to solve the regression problem, it uses a barcode matrix of lineage-defining mutations obtained from USHER and the mutation frequency and depth information to weight SNV frequency across each mutation site. These weights allow for prioritization of site-specific information as a function of sequencing depth which is ultimately used to generate a relative abundance of each of the known lineages.

Originally employed for abundance quantification of metagenomic transcripts, kallisto [26][27], has been repurposed to work with SARS-CoV-2 reads. Kallisto constructs an index from the RNA transcriptome with a de Bruijn graph that represents transcript k-mers. Reads are then pseudoaligned with the k-mers, and transcript abundance quantification is performed by likelihood function. For wastewater deconvolution, instead of using RNA-Seq transcripts, kallisto constructs the index of k-mer from multiple SARS-CoV-2 consensus sequences. The reads are pseudoaligned and estimated relative abundance are calculated.

LCS (Lineage deComposition for Sars-cov-2 pooled samples) is is a mixture model that determines SARS-CoV-2 variant composition in pooled samples by using a previously defined selection of mutations that characterize SARS-CoV-2 variants from publicly available sources and a matrix of variant signatures [28]. The matrix corresponds to the probability of finding an alternate sequence at any polymorphism from any variant. Along with the matrix, it uses the sequencing data containing counts of reads mapped to respective polymorphic loci. Minimap generated alignment files are then used for the estimation of the relative frequencies of the variants with maximum likelihood.

Kraken 2 [29] is a metagenomic sequence classification tool that uses alignments for taxonomic assignment. Similar to kallisto, Kraken 2 breaks down the sequences into k-mer and uses each of them to calculate a compact hash code to use as a query for finding the Lowest Common Ancestor (LCA) with a space seed searching scheme. This information is stored in a list and then used to form the classification tree where nodes are weighted based on the number of the k-mers linked with the taxon and root-to-leaf (RTL) path is weighted by addition of all the weights. The query sequence is then classified as the leaf to the maximum RTL path. For wastewater deconvolution, Kraken 2 uses the fastq sequences as the query and k-mer match to LCA for lineage identification and finally Bracken uses this lineage classification information to perform the relative abundance of sequence.

Freyja is frequently used as a component of other wastewater analysis pipelines, such as the C-WAP/Aquascope pipeline, which is a required step in the submission of wastewater sequencing results to NCBI and DCIPHER, the CDC reporting platform for wastewater monitoring. Kallisto, LCS and Kraken 2 are components of the C-WAP pipeline as well and obvious choices for inclusion in this analysis.

In addition to benchmarking the tools that are part of the C-WAP/Aquascope pipeline, we also evaluated four other commonly used lineage abundance estimation methods, as evidenced by the frequency of citations; LolliPop, Alcov, Lineagespot and VaQuERo. LolliPop [30] is a part of V-Pipe [31] and a part of processing viral sequencing data from NGS for the Swiss SARS-CoV-2 Sequencing Consortium (S3C) as a part of the SIB SARS-CoV-2 surveillance program. Lollipop solves the deconvolution problem by using a least square fitting approach and uses kernel-based smoothing to generate higher confidence relative abundance. It is designed to be integrated with Cojac [32] but can also be used independently. Alcov (Abundance Learning of SARS-CoV-2 Variants) treats lineage abundance estimation as an optimization problem using mutation frequencies [33]. Alcov focuses on nonsynonymous mutations only, and for each lineage-defining amino acid variant, the variant amino acid is back-translated into a nucleotide SNV at the appropriate genomic index. Lineage abundance estimation based on multiple variants is cast as an optimization problem using an ordinary least squares (OLS) approach, considering only mutations for which sufficient read depth is available. Lineagespot is another widely used deconvolution tool that detects variants and assigns lineages by the identification of mutational load by quantifying lineage abundance metrics, computing average allele frequency of all amino acid mutations and generates the mutational load as proportions [34]. Finally, we also considered VaQuERo, a relatively new method recently developed at CeMM (Center for Molecular Medicine), Vienna [11]. VaQuERo uses a SIMPLEX regression to deduce overall variant frequencies from the mutation patterns of individually selected variants. Both LolliPop and VaQuERo can use smoothing approaches to increase confidence in variant abundance estimates when presented with time series data, but the other tools are designed for the analysis of single samples.

As with any software tool that is designed to estimate an unknown, benchmarking and performance evaluation are essential for determining the most accurate software to identify mixtures of SARS-CoV-2 lineages in wastewater. Kayikcioglu et al, 2023 evaluated performance of 5 different deconvolution tools using simulated sequencing datasets [50]. Given that wastewater is a complex matrix, simulated data sets can be affected by assumptions about variant and error frequency that may not represent the behavior of nucleic acids in the wastewater matrix. These evaluations used the genomic simulator DeepSimulator, but the authors acknowledge that the simulation workflow did not take into account factors such as time, temperature, and the composition of the wastewater mixture. To account for the effect of these factors under standard wastewater processing conditions, we have sequenced synthetic RNA control mixtures. Controls were spiked into both water and complex wastewater extract backgrounds in known concentrations, and then sequenced on the Oxford Nanopore platform, to generate a gold standard dataset for benchmarking and evaluation of bioinformatic deconvolution tools. We illustrate what scenarios might determine which tool provides the most accurate results based on an expected input and identify strengths and weaknesses of available deconvolution tools, with the goal of informing future method development in wastewater analysis. The dataset is available through NCBI’s Sequence Read Archive (SRA) under the BioProject accession PRJNA1031245, and the protocol for preparation of control mixtures is available at <u>dx.doi.org/10.17504/protocols.io.261ged2jjv47/v1</u>, and analysis scripts and computational protocols are available at https://github.com/enviro-lab/benchmark-deconvolute.

## 2. Materials and Methods

Two types of controlled sample mixtures were assayed: controlled sample mixtures prepared in a buffer in isolation and controlled sample mixtures spiked into RNA extracts from wastewater samples. RNA extracts from both SARS-CoV-2 negative and SARS-Cov-2 positive samples were used as the matrix for spike-ins.

### 2.1 WW sample collection

Wastewater samples used as background for the constructed control mixtures, were collected as a part of the SARS-CoV-2 Campus monitoring program at the University of North Carolina Charlotte conducted by the Environmental Monitoring Lab [9]. The samples that were used in this project were collected in April 2023.

### 2.2 Concentration and RNA isolation

RNA concentration and extraction were performed using the KingFisher Flex with Nanotrap Microbiome A Particles, Nanotrap Enhancement Reagent, and the MagMAX Microbiome Ultra Nucleic Acid Isolation Kit. To make sure the samples are contamination free, nuclease-free water (NFW) was used as a negative control (known to be negative) in extraction and extracted alongside with the samples. Phosphate buffered Saline (PBS) was concentrated and extracted along with the samples as a processing control to make sure no contamination was introduced in the processing step. The automated Nanotrap® Microbiome A protocol provided by Ceres Nanosciences (2023) was used without modification.

### 2.3 SARS CoV2 detection by ddPCR assay

Extracted RNA was tested using droplet digital PCR (ddPCR) to quantitate SARS-CoV-2 viral RNA. Samples were tested using the CDC N2 primer and probe set with the BioRad one-step RT ddPCR Advanced kit [35]. A positive control and NFW negative control were included in the assay. If out of 10000 droplets at least 3 are positive, the sample was then considered as positive. All the results were analyzed with the QuantaSoft™ Software, Regulatory Edition #1864011 [36]. The reported average concentration of SARS-CoV-2 in the positive wastewater background samples was 2.23 copies/ul.

### 2.4 Assay-ready synthetic mixed control preparation

Assay-ready synthetic RNA controls representing various SARS-CoV-2 major variants were sourced from Twist BioSciences (San Francisco, CA). These controls were used in known concentrations and in a variety of combinations of variants and mixture complexities. 15 different synthetic controls were used - Control 15 (Alpha-103909), Control 17 (Gamma-104044), Control 23 (Delta-104533), Control 48 (Omicron - BA.1 lineage-105204), Control 51 (B.1.1.529+BA.2-England-105346), Control 2 (Wuhan hu-1 from china-102024), Control 6 (Wuhan hu-1 from California-102918), Control 50 (B.1.1.529+BA.2-Australia-105345), Control 64 (BA.5-England-106196), Control 62 (B.2.12.1-Denmark-105865), Control 66 (BA.4-Texas-106198), Control 67 (BA.4-California-106199), Control 63 (B.2.12.1-USA-105857), Control 65 (BA.5-USA-106197) and Control 19 (Iota-104529). From these 15 controls, 38 different mixtures were prepared, as outlined in Supplemental Table 1.

### 2.5 Spike-in mixed control preparation

All mixed controls were also spiked into RNA extracts from SARS-CoV-2 positive and SARS-CoV-2 negative wastewater samples, to provide a more realistic nucleic acid matrix for subsequent amplification and sequencing steps. Wastewater samples collected in April 2023 (04/13/23-04/27/23) were quantified as described above. The ratio of synthetic controls in the mixed control in water background was maintained to allow for comparison between control mixtures and corresponding mixtures in wastewater backgrounds.

### 2.6 Sequencing control mixtures

Whole genome sequencing based on tiled amplicon amplification was performed with two different primer sets - ARTIC v4.1 primer, which generates 400bp long amplicons and VarSkip short v2a, which generates 550 bp long amplicons. For both of the reactions, the NEBNext^®^ ARTIC SARS-CoV-2 Companion Kit (New England Biolabs) was used. ARTIC v4.1 primers from IDT were used in place of the ARTIC v3 primers that are contained in the NEBNext kit in a 1:100 dilution. For ARTIC, the protocol was followed as outlined in the NEBNext ARTIC instruction manual (E7660) and for Varskip the protocol was followed as outlined by Ramachandran et al. [18]. For barcoding, the Native Barcode Expansion Kit (Exp-NBD196) from ONT was used. Samples were sequenced on the Oxford Nanopore PromethION using R9 flow cells.

### 2.7 Sequence trimming and filtering

Samples were analyzed via an in-house covid-analysis pipeline [37] which trims and filters reads using artic, guppyplex, Kraken 2, and Porechop before running through the artic minion command of the ARTIC pipeline (a bioinformatic pipeline that is articulated to analyze nanopore sequencing data generated from tiling amplicon schemes) [38]. Samples were also analyzed with the C-WAP pipeline [39] which was used during the COVID-19 pandemic for analysis of sequencing data from wastewater samples by the CDC national wastewater network. For the ARTIC pipeline, the sequence length cutoff range was 305-505 bp on sequences generated using the ARTIC 4.1 primers and for Varskip, it was 475-675 bp. The .bam, .fastq, or .vcf files generated by the covid-analysis pipeline were then used as input for all subsequent analyses, depending on the requirement of each deconvolution tool.

**Figure 1.**
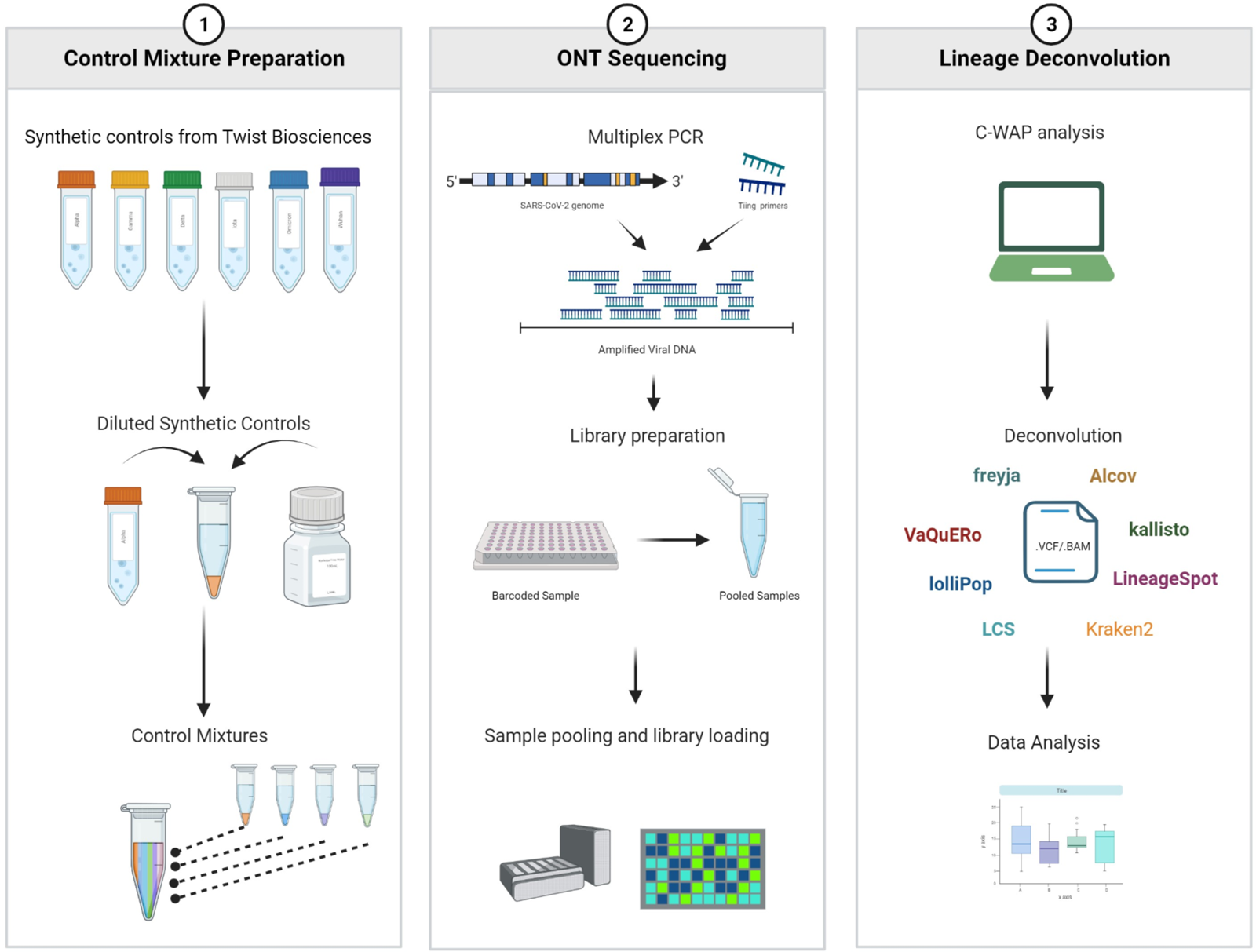
Overview of SARS-CoV-2 control mixture preparation, sequencing and computational analysis

### 2.8 Deconvolution of mixtures

Freyja [40], kallisto [26], LCS [28] and Kraken2/Bracken [29] were selected due to their use in the CFSAN’S C-WAP pipeline and LolliPop [30] was selected for its use in V-pipe. In addition to these, LineageSpot [34], Alcov [33], and VaQuERo [11] were selected to look at different and newer approaches to deconvolution. The project github repository linked in Supplemental Materials 1 provides analysis scripts used in the project along with a complete description of parameter and option choices used with each tool.

## 3. Results and Discussion

Deconvolution of SARS-CoV-2 lineage abundance from wastewater extract sequencing data can be accomplished using a variety of algorithms. The deconvolution methods assessed in this paper can be grouped depending upon a variety of methodological choices, including whether classification is based on recruitment of reads to a reference genome or on a match of detected SNVs to a pattern of defining mutations; the source of the reference genome or reference lineage-defining mutation information used for classification; the algorithmic approach used; and whether results are reported as relative abundance of an entire lineage, or as proportion of individual lineage-defining SNVs detected. There are also operational differences in each computational workflow, especially as to whether the input required is the raw .fastq file, a pre-aligned .bam or .sam file, or a .vcf file extracted from a read alignment (Table 1). Definition of variants by each method is with respect to the original Wuhan strain of SARS-CoV-2, except for kallisto, which uses recruitment of reads to a collection of reference strain genomes.

**Table 1:** Comparison of Deconvolution Tools.

| Tool | Algorithm | Reference set source(s) | Reference Reconstruction | Input | Output |
| --- | --- | --- | --- | --- | --- |
| Alcov | Optimization | outbreak.info + covvariants.org | Mutation based | .bam | Relative abundance |
| Freyja | Depth weighted least absolute regression | cov-lineage | Mutation based | .bam, ref-fasta | Relative abundance |
| kallisto | Pseudoalignment of hashed k-mer | GISAID or UShER (UCSC) | Reference based | fastq, ref-fasta | Relative abundance |
| LCS | Statistical regression Problem | GISAID or WHO | Mutation based | fastq | Relative abundance |
| LineageSpot | Calculation of allele frequency of unique and AA mutations | outbreak.info | Mutation based | .vcf | Mutation proportions |
| LolliPop | Least square problem | cov-spectrum.org or covariants.org or [PHE Genomic's Standardised Variant Definitions](https://phe.genomic.org) | Mutation based | .bam | Relative abundance |
| VaQuERo | SIMPLEX regression | GISAID and ECDC | Mutation based | .bam or .vcf | Relative abundance |
| Kraken 2 | K-mer match to LCA taxa | GISAID | Reference based | .fastq | Relative abundance |

### 3.1 The wastewater sample background changes the properties of sequence data

3.2 Wastewater sample processing can result in biases in downstream quantitation or sequencing processes due to sample quality and sample chemistry. The choice of extraction methods may result in fragmentation of the input RNA [41], leading to shorter sequence reads of lower quality, and the chemical makeup of extracted samples may include small molecules that inhibit the amplification steps in detection [42][9][43] and sequencing. To understand the impact of the wastewater extract matrix itself on sequencing outcomes, we compared sequencing results generated for control mixtures spiked into water background (WB), SARS-CoV-2 negative wastewater RNA extract background (NWRB) and SARS-CoV-2 positive wastewater RNA extract background (PWRB). We compared the percent genomic coverage and total read count generated from each sample. Figure 2a shows percent coverage across the genome for each sample. Mean coverage for WB (mean=96.925, std. dev.=0.763) differed significantly from NWRB (mean=95.580, std. dev.=1.136), and PWRB (mean=95.510, std. dev.=2.010), as determined by one-way ANOVA (F=12.252, p<1e-4). Tukey’s HSD test showed significant differences between WB and each wastewater background (p<0.01 for each), but the mean coverage values for samples in wastewater backgrounds did not vary significantly from each other (p>0.01). Read counts showed significant differences between each of WB (mean=73538.6, std. dev.=1214.6), NWRB (mean=70281.7, std. dev.=1974.8), and PWRB (mean=72270.6, std. dev.=1574.5) with Artic v4.1 primers, as determined by one-way ANOVA (F=39.119, p<1e-12) and Tukey’s HSD test (p<0.01 between each background). Normalized read counts for each background and both primer sets can be seen in Figure 2b. This suggests that the complex matrix of wastewater RNA extract impacts sequencing coverage whether or not additional SARS-CoV-2 RNA is present in the sample. Although a tiling amplicon genome sequencing approach is typically expected to provide uniform and complete coverage of the target genome [44], the complexity of the wastewater matrix clearly reduces this coverage. This is also reflected in the comparison of normalized read counts. The significant difference between each group’s read counts points to the fact that introduction of different backgrounds led to failure of uniform mapping and inconsistent coverage. Despite having the same reaction conditions, PWRB and NWRB showed significant differences in read counts. This suggests that the sample heterogeneity and compounds carried through into the wastewater RNA extract during processing can impact the successful mapping of reads.

**Figure 2.**
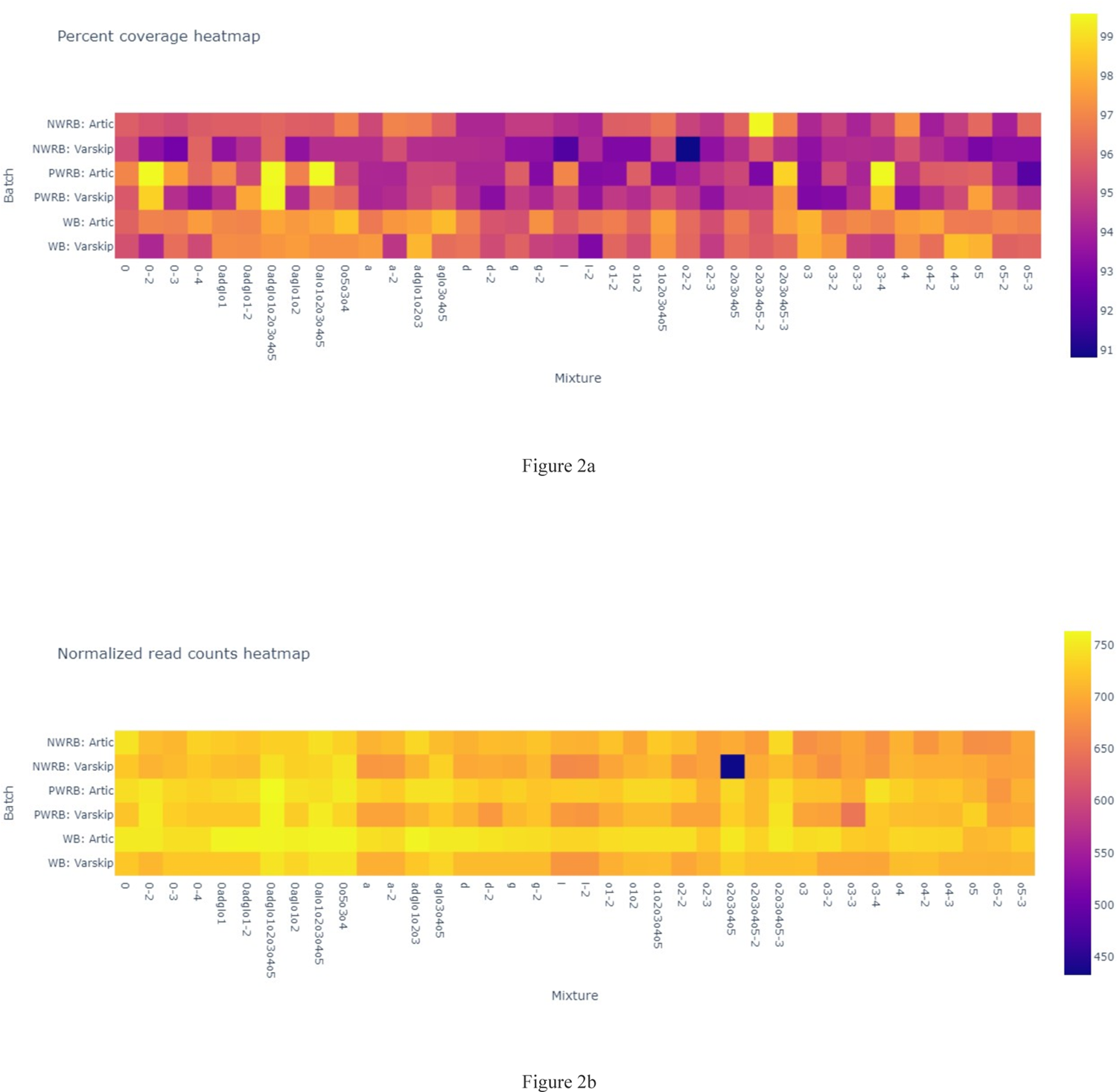
Overview of sequencing data quality with different parameters - 2a) Sequencing data quality based on percent coverage, 2b) Sequencing data quality based on read counts

### 3.3 The influence of tiling amplification scheme on sequence coverage is significant

For wastewater amplification there are numerous primer choices available, and the amplification scheme may affect the outcome of sequencing in different ways [45]. To evaluate the effect of the primer scheme, we replicated our experiments using two different tiling primer sets: Artic v4.1 and VarSkip short v2a. The analysis is shown in Figure 2a. Mean coverage and read counts obtained from spike-ins in WB showed significant differences from those obtained from spike-ins in NWRB and PWRB with Artic v4.1, as outlined in section 3.1. For Varskip, all three groups varied significantly from each other based on one-way ANOVA (F=26.194, p<1e-9) and Tukey’s HSD test (p<0.01 between all groups), with mean percent coverage the lowest for NWRB (mean=94.084, std. dev.=1.074), increasing with PWRB (mean=95.168, std. dev.=1.590), and increasing again with WB (mean=96.264, std. dev.=1.219). This is similar to the pattern of coverage values obtained using the Artic v4.1 primers, and continues to suggest that the wastewater matrix impacts the quality of sequence data produced regardless of the primer scheme chosen. Read counts did not show statistically significant differences between backgrounds (one-way ANOVA: F=3.433, p>0.01, WB_mean_=52889.9, WB_std. dev_=1151.4, NWRB_mean_=51603.6, NWRB_std. dev_.=3543.9, PWRB_mean_=52761.1, PWRB_std. dev._=1667.5). However, comparing coverage attained by Artic v4.1 (mean=96.01, std. dev.=1.538) and Varskip v2a (mean=95.17, std. dev.=1.578), the two primer schemes showed significantly different coverage, t(78)=4.036, p<0.001 with Artic having the higher mean and lower standard deviation. This suggests that use of the Varskip primer scheme reduced amplification efficiency in a small but significant way. Even though some specific primer pairs may vary in amplification efficiency depending on the circulating lineage in the sample, the Artic v4.1 primer scheme showed more amplification efficiency with these samples regardless of the sample matrix. Deconvolution method comparisons are therefore based on the Artic v4.1 data, with corresponding results for the Varskip amplification included in as Supplemental Figures 1, 2 and 3.

### 3.3. There are significant differences in variant identification between deconvolution tools

To assess the performance of the chosen deconvolution methods, we first compared the ability of each of the methods to detect the strains included in the original sample and in the expected proportions. Median pairwise L2 abundance norms, calculated using the approach described in Ye et al [46] provides a summary of the accuracy of abundance estimation across methods, offering a comparison between expected vs estimated abundance, and also among the tools themselves. Apart from Kraken2, all the methods examined predicted presence and relative abundance of the expected lineages with high accuracy (Figure 3). The predictions between the tools were not statistically significant, again with the exception of kraken and sometimes kallisto, as determined by one-way ANOVA (F=10.655, p<1e-11) and Tukey’s post hoc HSD Test. In Supplementary Table 1, we show the expected and predicted composition for each sample mixture. All of the deconvolution methods tested in this study could identify the major lineages that were present in the control mixtures, except for Kraken2 (Supplemental Figure 4).

While most of the methods identify the variant in the mixed control dataset at the lineage level, there are misidentifications in the sub-lineage level of the variants. Most of these tools use a mutation-based reference set to define lineages and sublineages. As SARS-CoV-2 is a pathogen with a high mutation rate, the presence or absence of a single significant lineage-defining mutation can cause misidentification of sub-lineages. For example, BA.4 and BA.5 share more than 50 similar mutations [47] and BA.1 and BA.2 share 29 similar mutations [48]. At this degree of similarity, a single missing lineage-defining SNP has the potential to result in a misidentification. This is compounded by the fact that lineages are being defined and classified based upon amplicons and may not be able to be easily assigned to one variant.

Misidentification was also seen in older variants such as Iota, because some of the tools did not incorporate mutations for variants that were not considered as variants of concern or interest, such as Iota. Most of the deconvolution tools also did not explicitly identify the original Wuhan-hu-1 strain when it was a component of the mixture, other than Freyja. As most of these tools define lineages based on mutations relative to the Wuhan-hu-1 strain, and there is no mutation present in Wuhan-hu-1 relative to itself, none of them label this lineage correctly when it is present in a wastewater mixture, perhaps because its occurrence is a circumstance that is now unlikely other than in archival samples or constructed standard mixtures like those created for this study. VaQuERo, if it is unable to detect any lineage or sublineage based on mutations, will identify that sample as Wuhan-hu-1 by default.

**Figure 3:**
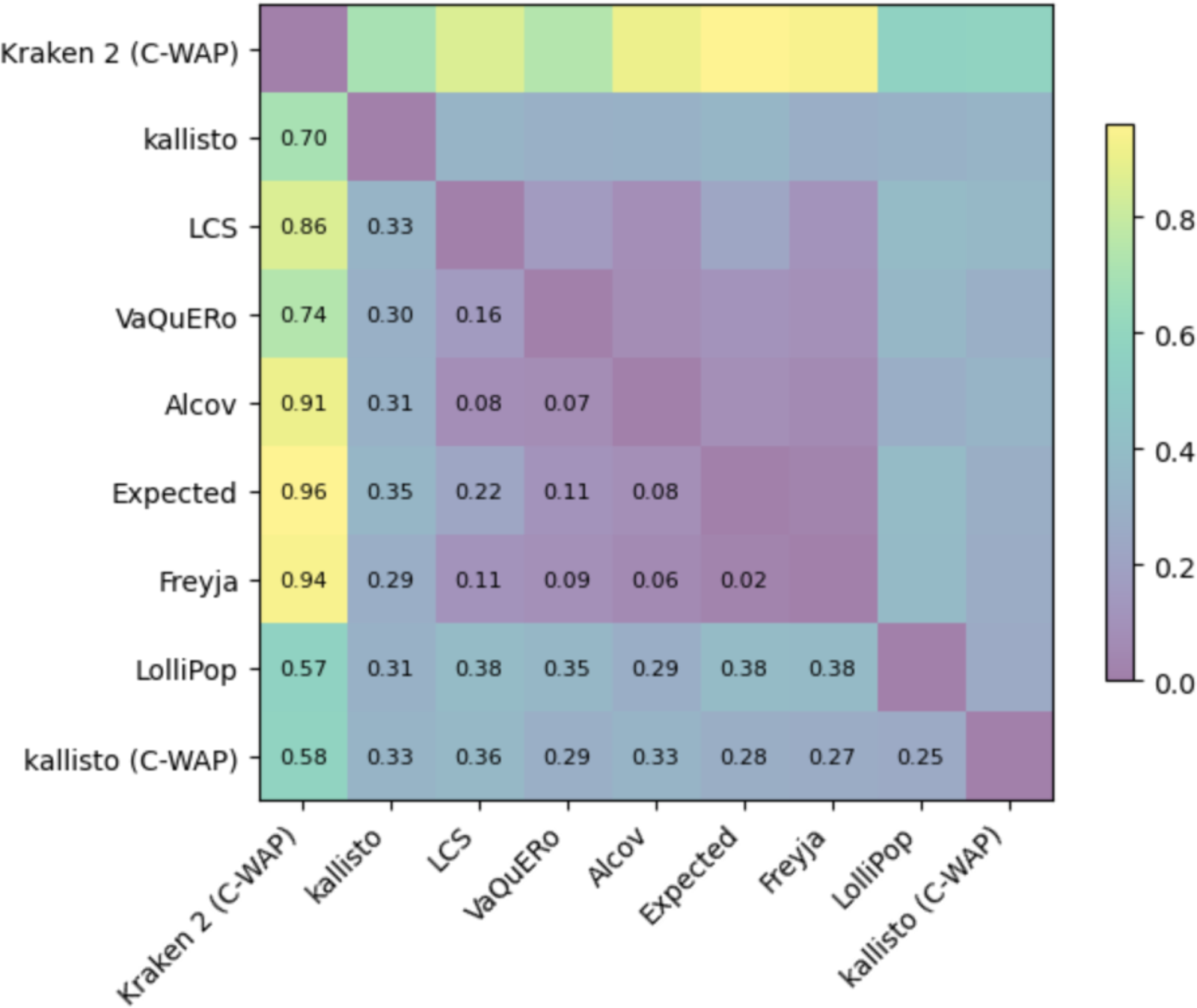
Median pairwise L2 abundance norms between deconvolution tools

### 3.4. The wastewater background had only subtle effects on abundance estimation

In section 3.1, we showed that the composition of the sample matrix affects the coverage of sequencing. We also tested whether the sample matrix of the controls that are spiked into affects the estimation of relative abundance of lineage. We found that the estimation of the relative abundance of lineages changes very little regardless of whether it is spiked into negative or positive wastewater. The variances between samples from different backgrounds were insignificant (p>0.05 for all) via ANOVA performed to test the effect of sample matrix on the estimated relative abundance for all abundance-estimating tools. The small change in the estimated abundance can be attributed to the extra complexity of a metagenomic sample and the effect of addition of lineages of SARS-CoV-2 from the positive wastewater background to the synthetic control mixtures, but this does not negatively impact the performance of the tools (Figure 4). This could also be showing that the deconvolution tools are not extremely sensitive to the coverage and sample matrix for relative abundance estimation.

For the small differences that were observed, ANOVAs were performed for each lineage to see how those differences varied depending on the water or wastewater background of a sample. These results are shown in Table 4. The lineages that preceded Omicron tended to show statistically signficant differences between samples from wastewater backgrounds and water backgrounds, suggesting that sample background affects the lineage assignment of older variants more significantly than the newer ones. The addition of wastewater, which has a complex composition of inhibitors and carry through compounds, can inhibit the PCR and sequencing reactions and affect the sensitivity of variant detection. Additionally, the absence of defining mutations in the reference lists of deconvolution tools, for older/non-variant-of-interest lineages (as we have seen in Section 3.3) can cause misidentification and misestimation.

**Figure 4:**
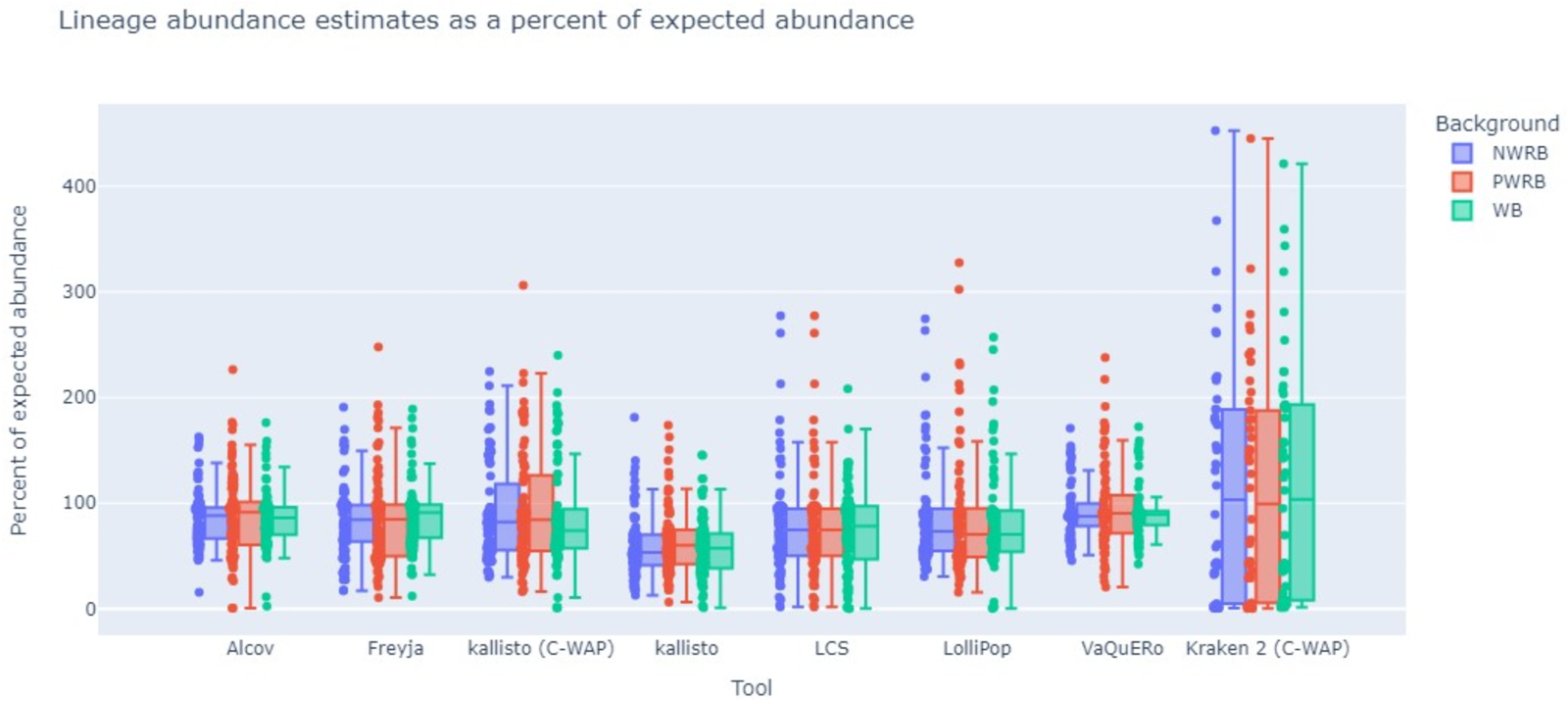
Comparison of relative abundance of datasets in different backgrounds

**Table 4.**
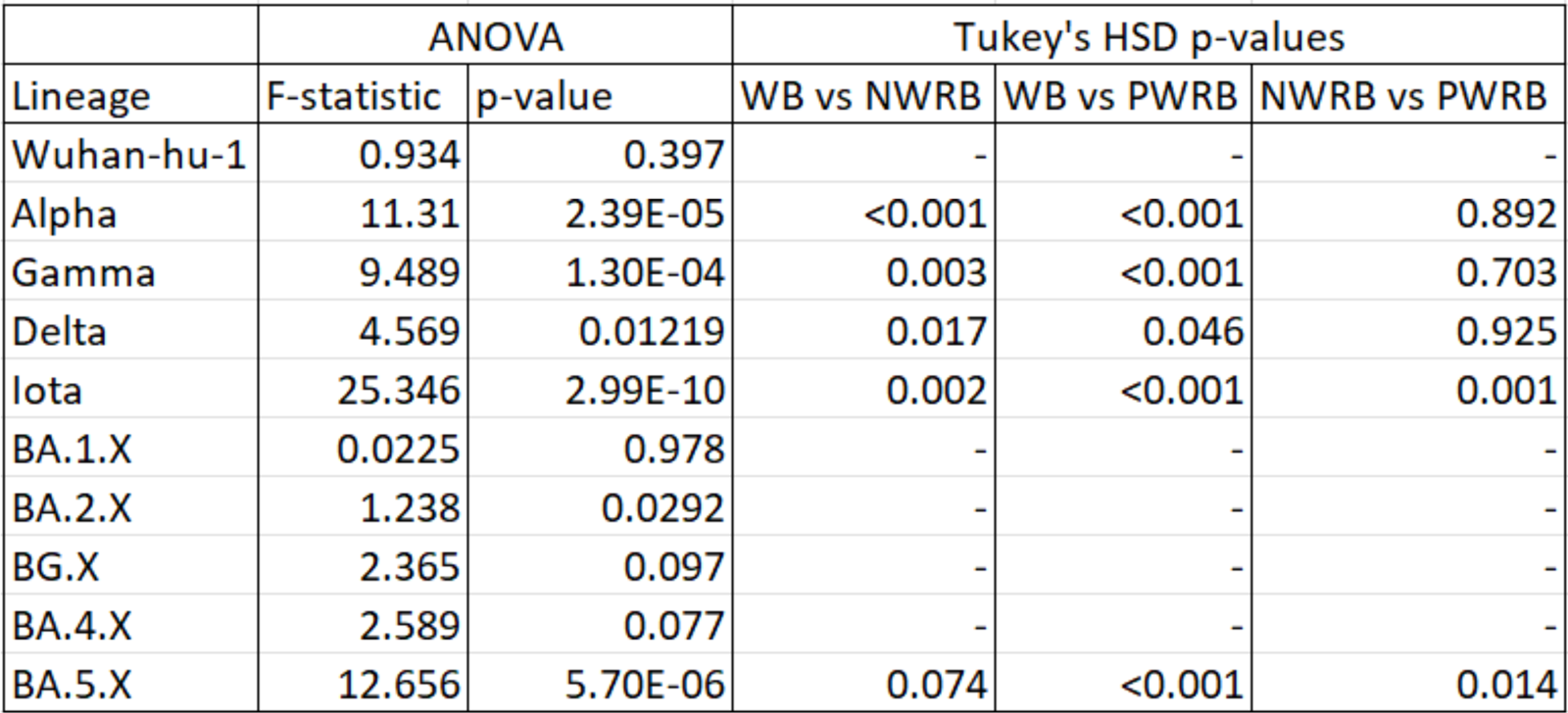
ANOVA between backgrounds and Tukey’s HSD results when grouped by lineage.

| Lineage | ANOVA |  | Tukey's HSD p-values |  |  |
| --- | --- | --- | --- | --- | --- |
|  | F-statistic | p-value | WB vs NWRB | WB vs PWRB | NWRB vs PWRB |
| Wuhan-hu-1 | 0.934 | 0.397 | - | - | - |
| Alpha | 11.31 | 2.39E-05 | <0.001 | <0.001 | 0.892 |
| Gamma | 9.489 | 1.30E-04 | 0.003 | <0.001 | 0.703 |
| Delta | 4.569 | 0.01219 | 0.017 | 0.046 | 0.925 |
| Iota | 25.346 | 2.99E-10 | 0.002 | <0.001 | 0.001 |
| BA.1.X | 0.0225 | 0.978 | - | - | - |
| BA.2.X | 1.238 | 0.0292 | - | - | - |
| BG.X | 2.365 | 0.097 | - | - | - |
| BA.4.X | 2.589 | 0.077 | - | - | - |
| BA.5.X | 12.656 | 5.70E-06 | 0.074 | <0.001 | 0.014 |

### 3.5. Error types varied significantly between deconvolution methods

To understand how the lineage abundance estimated by each method differed from the expected abundance, the estimated relative abundance in each control dataset with different backgrounds were analyzed with respect to the percent abundance of lineages. (Figures 5a, 5b, 5c). As seen in Figure 5a, certain tools underestimated lineage abundance more often with certain lineages. LolliPop, Vaquero and LCS all underestimated the abundance of the older variants, mostly Delta and Iota; though LolliPop underestimated most of the lineages. Among the pre-Omicron variants only, Alpha was overestimated in some of the mixtures by Freyja and Alcov. Gamma was the only variant that had similar relative abundance estimated by all the tools. Conversely, the abundance of Omicron and its sub lineages are often overestimated by most of the deconvolution tools. Kallisto’s performance varies greatly depending on the reference database chosen; this is discussed further in section 3.7.

When considering the effect of the wastewater background (Figure 5b and 5c), some patterns emerge; Alpha, Delta, and Iota are often slightly underestimated in negative and positive wastewater backgrounds. This underestimation might be due to inhibitors that can be carried through the RNA extracts from the processing step and affect sequencing and subsequent lineage detection sensitivity. The sub lineages of Omicron are overestimated in the positive wastewater background relative to the other two backgrounds. This is expected as positive samples were collected from April 2023 when most circulating lineages are Omicron, leading to an increase in Omicron concentrations in the samples.

**Figure 5:**
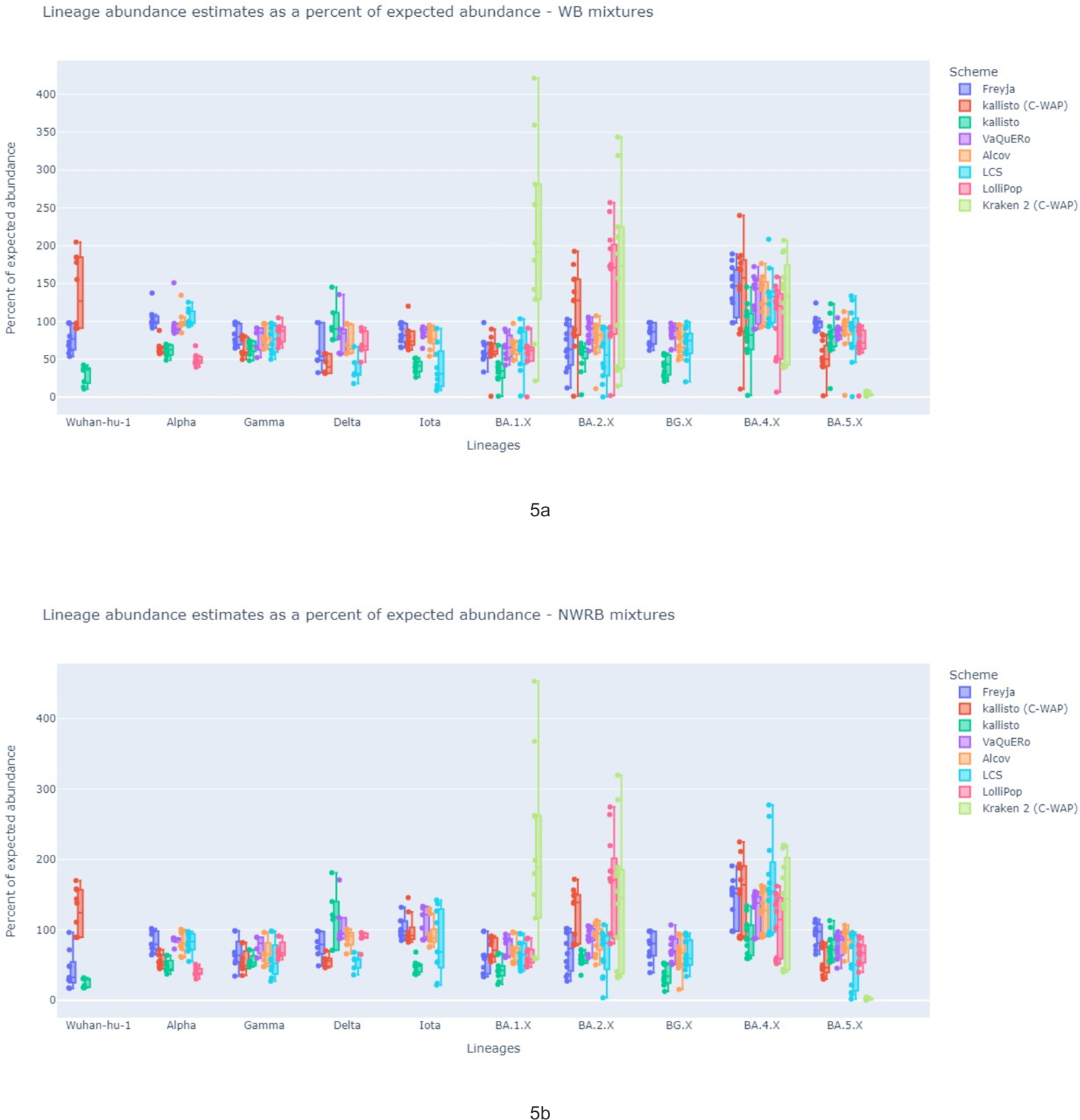
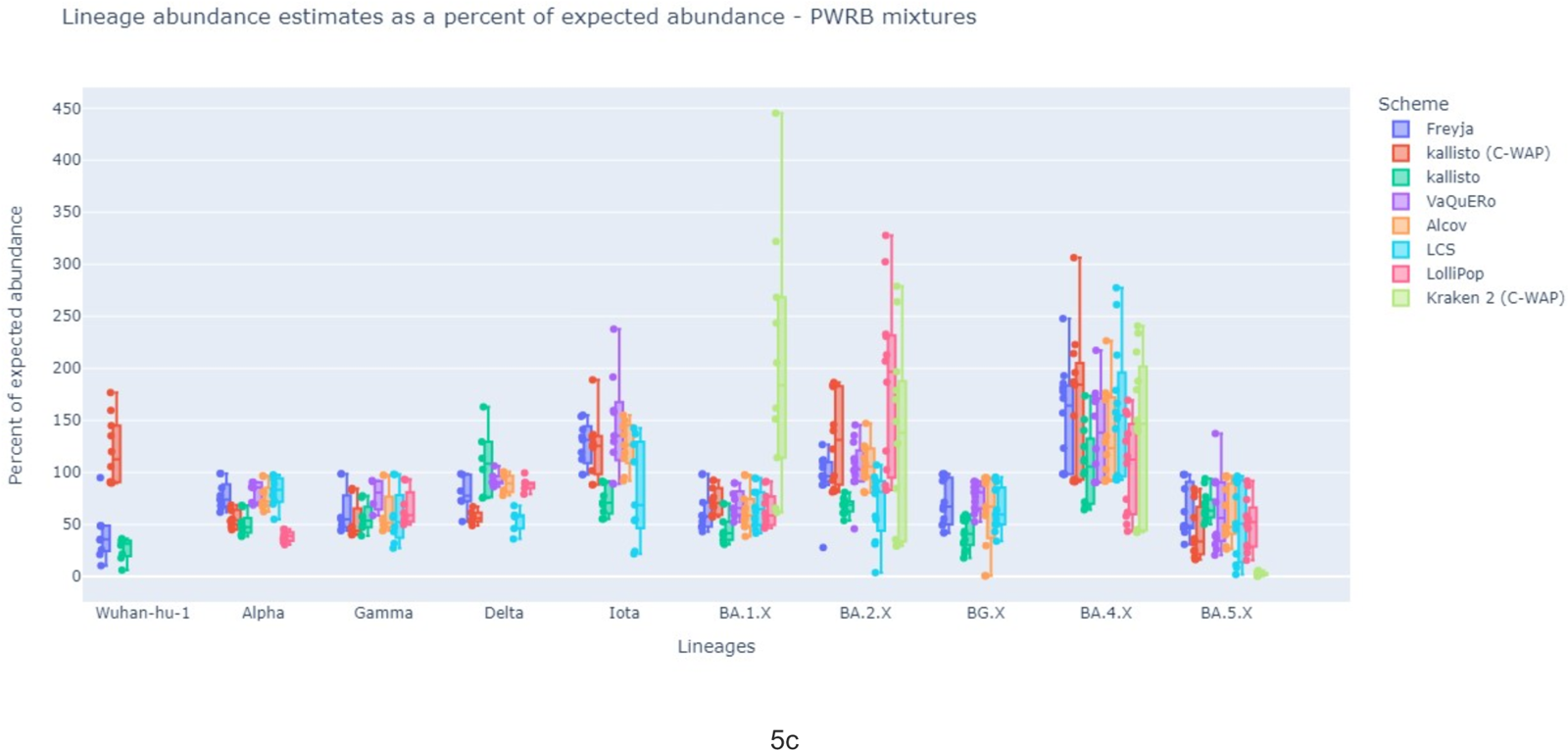
Overview of change in expected abundance of lineages along with the change in the environment of control mixtures

### 3.6 Freyja yielded the most accurate lineage compositions and fewest false negatives

**3.7** One of the main objectives of this study was to identify which deconvolution tool most accurately deconvolutes samples of known composition, in terms of overall accuracy and false positive or false negative identifications. Output of all methods tested was compared to the expected relative abundance of strains in the prepared samples. Results of each method were also compared to one another. We found that Freyja outperformed other tools in both accuracy and fraction of false negatives.

The median pairwise L2 abundance norms calculated in Figure 3, generated from the distances between expected control vector and deconvolution tool datasets, shows that Freyja has the lowest distance (0.02) from the expected vector, indicating that Freyja has the highest accuracy in identifying the relative abundance of lineages. Supplementary Figure 1 shows Freyja’s detection of lineage composition in comparison to other deconvolution tools when the Varskip primer set is used. Similarly, the lowest distance is also Freyja to expected with 0.03. When considering false positives, Freyja consistently identified the lineages with minimal false positives and zero false negatives (Figure 6a, 6b). With highest accuracy, no false negatives and few false positives, Freyja most accurately replicates the lineage identification and abundance of expected control mixtures. The expanded lineage lineage detection composition for the other tools tested can be found in Supplemental Figures 5.

When comparing the effectiveness of each tools’ lineage detection, it was found that VaQuERo closely followed Freyja in performance. The rest of the tools detected multiple false positives, lineages which were not present in the control mixture dataset and sometimes also failed to detect the spiked-in lineages correctly. As false positive identification presents a major challenge in wastewater lineage deconvolution, the relatively low false positive calls in Freyja and VaQuERo carry significant weight in tool choice. When we consider how each algorithm works, Freyja uses each of the mutations from the alignment files and their respective sequencing depth to perform the depth weighted least absolute deviation regression and VaQuERo also uses all unique and non unique marker mutations to solve the deconvolution problem using SIMPLEX regression approach. This inclusivity of all the mutations while estimating the lineage composition might be the key to their better performance in false positive identification.

**Figure 6a:**
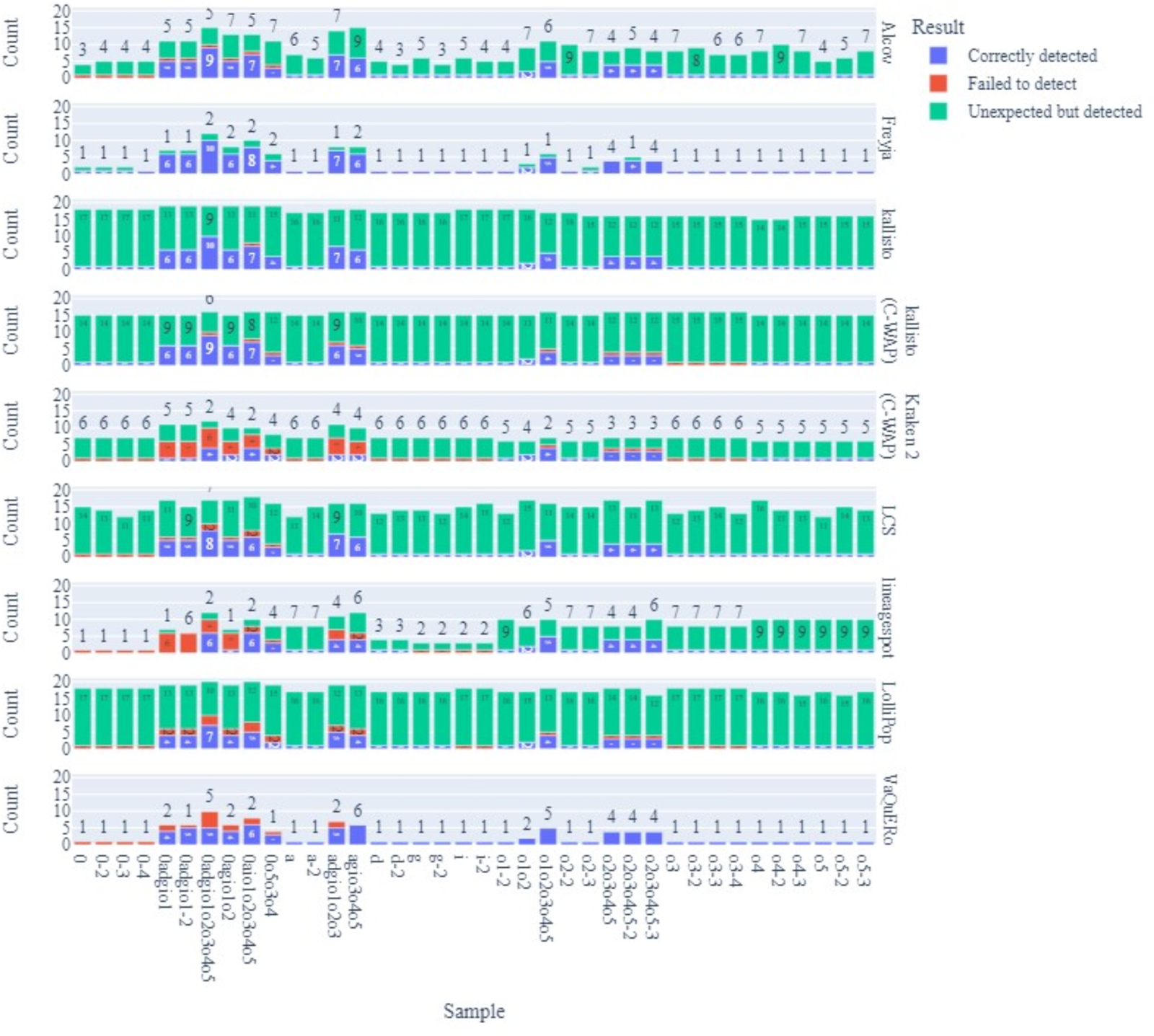
Detection of expected lineage by the deconvolution tools (WB mixtures)

**Figure 6b:**
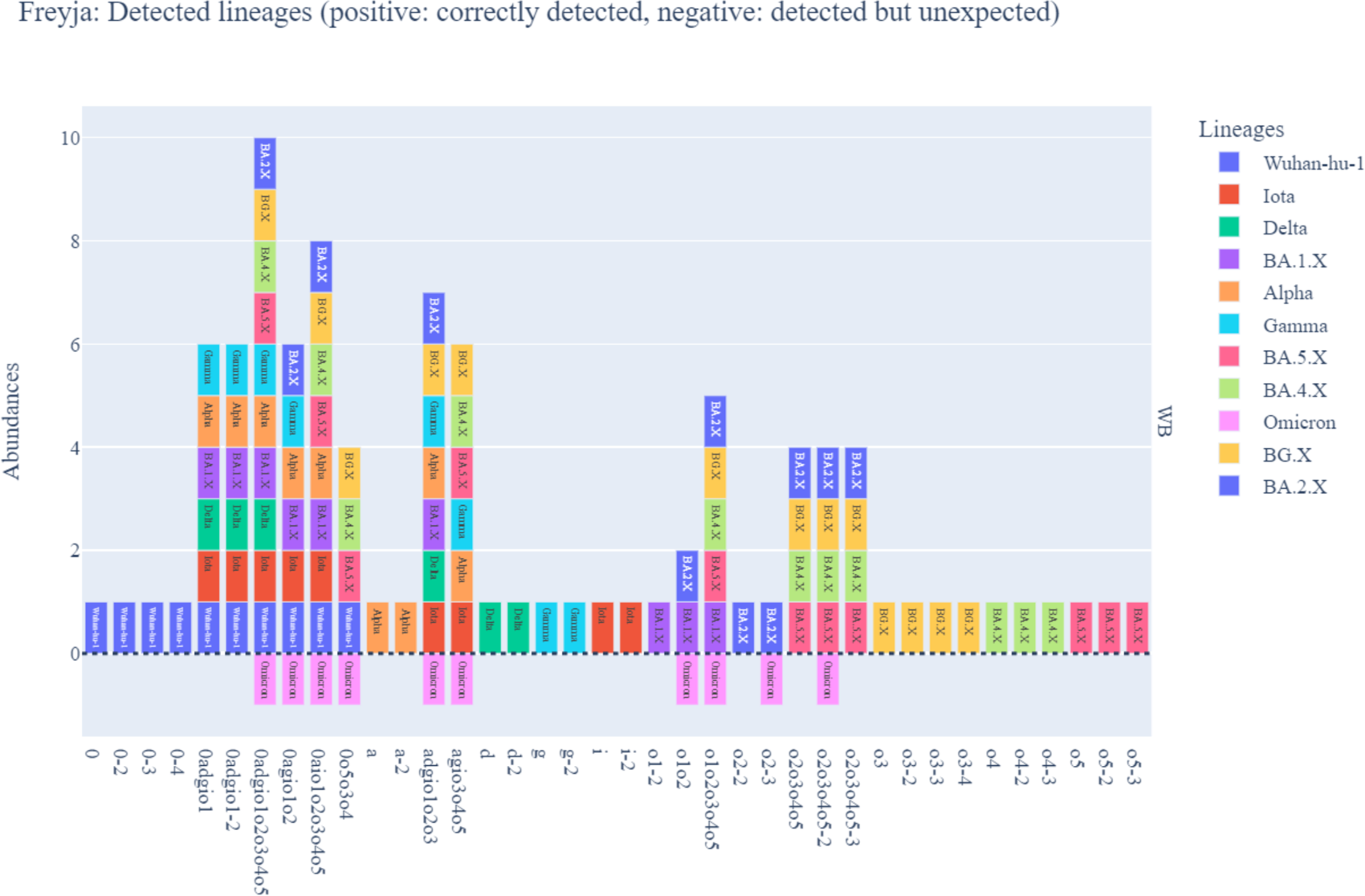
Figure 6b: Detected lineage composition by Freyja (WB mixtures)

### 3.7 Kallisto performance was strongly dependent on the reference database

Kallisto is originally a RNA transcript quantification tool. As the variant abundance estimation is found to be computationally similar to RNA transcript abundance estimation [27], it has been repurposed to be used as a wastewater deconvolution tool for relative abundance estimation of the SARS-CoV-2 variants. Kallisto takes a set of reference sequences that contains multiple genomic sequences per lineage, with the recommendation to include multiple sequences per lineages to reduce biases relating to within-lineage variation [27]. As a consequence, the composition of the reference set can significantly affect the performance of a reference based deconvolution tool [49]. We developed two databases to test how the performance of kallisto varies with respect to the database used.

The in-house database preparation is described in the configuration parameters (Supplemental Materials 1) and includes 406 sequences with up to four sequences per lineage for 299 different lineages. On the other hand, the C-WAP databases for both kallisto and Kraken 2 used 30 sequences with up to six sequences for each of the following 13 included lineages: Wuhan-Hu, Alpha, Beta, Gamma, Eta, Epsilon, Delta, Iota, Kappa, BA.1, BA.1.1, BA.2, and BA.3 [50]. Those databases were produced one time, receiving no further updates over time. The in-house database used lineages that were more specific than those in the C-WAP database, including many sublineages of the lineages listed for C-WAP’s database. Figure 3 shows that the accuracy of kallisto varies between these reference databases. Kallisto run with the C-WAP database has the median distance of 0.28 with respect to the expected abundances, and kallisto with the in-house database has a distance of 0.35. This means that the accuracy of kallisto is improved when using the C-WAP database. Similarly, more false positive lineages were identified with the in-house database (Figure 6a). We hypothesize that because the in-house database has significantly more reference sequences per lineage, the algorithm may have had difficulty with unambiguous read assignment and aggregation into lineages. Therefore, the goal of correctly interpreting within-strain variation may be at odds with the goal of unambiguous read mapping, and the reference database should be chosen with care.

### 3.8 Interpretation of increasingly complex variant profiles is a future challenge

The reference sequences necessary for deconvoluting wastewater mixtures rely on the availability of clinical sequencing of SARS-CoV-2 positive patient samples. As PCR-based lab testing has declined in favor of at-home rapid testing, access to clinical specimens has decreased significantly. This affects the availability of reference sequences for current circulating variants. Theoretically, the drift of current sequences away from the abundant reference data available for the 2020-2022 time period at some point starts to impact the performance of deconvolution tools in identifying lineages. If this is indeed happening, we should see an increase in the abundance of unidentified or unclassified lineages in wastewater across time.

With the continuously growing number of variant sublineages, an abundance threshold is often applied to filter out very low abundance lineages and simplify visualizations. As part of our research program, we have been sequencing wastewater samples across North Carolina using the ARTIC primer set and Freyja as part of C-WAP for deconvolution. If we examine data from September 2022 to July 2023 (Figure 7), we can see that the trendline shows abundance of “unclassified by Freyja” and “below abundance threshold” lineages increasing over time. The number of circulating lineages in wastewater samples that fall below typical abundance thresholds is continuously increasing (Figure 7), and without a threshold the visualization and analysis of wastewater will lose its clarity and interpretability of current trends. It is also creating a challenge for the deconvolution algorithms to identify the variants with accuracy. We can see in 3.6 that the most accurate output generating tool Freyja is overcalling the generic Omicron mostly in the complex mixtures with multiple controls in varying proportions starting from 625 - 2500 copies/ul. We hypothesize that with this continuously growing variant abundances needs to be taken into account to improve the algorithm performances . These trends also suggest that other algorithms and methods to deconvolute wastewater samples are going to need to be developed that do not strictly rely on existing reference genomes and variant calls. Studies that can predict surge of infection can be informational in this scenario to help with public health measures to clinical sequencing as well as wastewater. This increasing growth of unidentified lineages supports the concern that unidentified sequences are going to continue to increase as we have fewer reference genomes for comparison, calling into concern the ability of these deconvolution tools to accurately identify circulating lineages.

**Figure 7:**
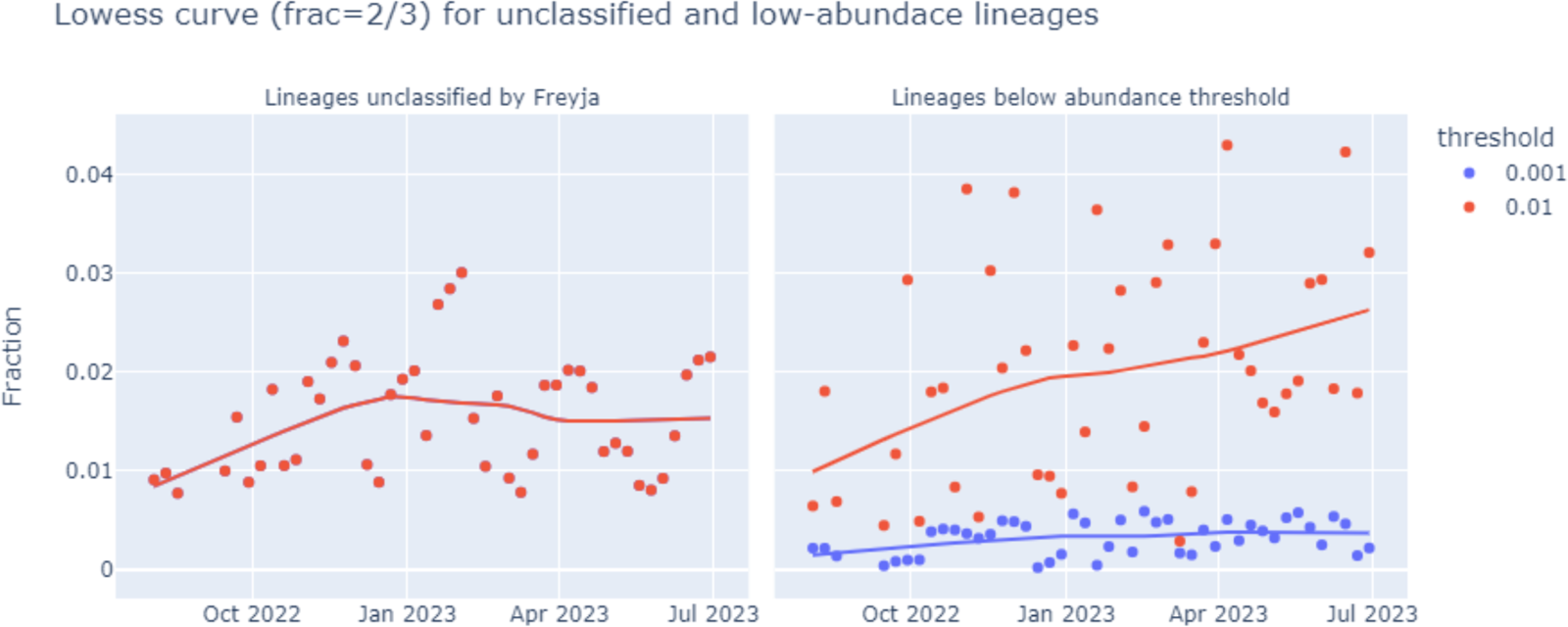
LOWESS trendline on NCDHHS wastewater sequencing data

## 4. Conclusions and Future Work

This study addresses the ability of commonly used deconvolution methods to distinguish the presence and abundance of SARS-CoV-2 variant spike ins when controlled mixtures are sequenced. We find that Freyja, which has been widely adopted throughout the COVID-19 pandemic, produces variant abundance calls with the closest relationship to the expected ratios when tested on controlled variant spike-in mixtures. The error profile produced by Freyja is dominated by false positive lineage identifications rather than false negatives, including the identification of indeterminate omicron lineages that do not correspond to any particular control, which are seen mainly in complex mixtures containing multiple spiked in variants. We also examine the influence of the wastewater background on deconvolution outcomes. We find that the impact of the wastewater matrix on variant deconvolution is insignificant, despite significant differences in sequence coverage resulting from the wastewater matrix. The influence of different tiling amplicon primer schemes on deconvolution outcomes is also negligible. The impact of the concentration and extraction process on viral RNA detection and quantitation has been extensively studied, and the impact of that process on wastewater variant sequencing could be investigated by repeating this study beginning from encapsulated viral controls spiked into raw wastewater, but that is a separate question, less about bioinformatics method performance than about sample processing and chemistry. It would likely be interesting to repeat this study on the commonly-used Illumina sequencing platform as well. However, given that we observe minimal differences in variant identification due to either tiling amplification scheme or exposure of RNA to the wastewater extract background, and the ongoing improvements to basecalling error rates on the Oxford Nanopore platform, the influence of the sequencing platform on variant deconvolution is likely to be relatively subtle as well.

## Data Availability

All data produced are available online at

https://dx.doi.org/10.17504/protocols.io.261ged2jjv47/v1

https://github.com/enviro-lab/benchmark-deconvolute

## Acknowledgements

We gratefully acknowledge all data contributors, i.e The Authors and their Originating laboratories responsible for obtaining the specimens, and their Submitting laboratories for generating the genetic sequence and metadata and sharing via the GISAID Initiative, on which this research is based. The authors of a few tools were most helpful when asked for clarification on how to use their tools or adapt them to the constraints of this study. We would like to thank Fabian Amman and his team from VaQuERo, Art Poon from Gromstole, and Ivan Topolsky from LolliPop for their assistance. We would also like to acknowledge BioRender, which was used to generate the Graphical abstract and the overview figure. Funding for this project was provided by the North Carolina Department of Health and Human Services.

## Abbreviations

SARS: Severe Acute Respiratory Syndrome
COVID-19: Coronavirus Disease 2019
WBE: wastewater-based epidemiology
WB: water background
NWRB: SARS-CoV-2 negative wastewater RNA extract background
PWRB: SARS-CoV-2 positive wastewater RNA extract background
NWSS: National Wastewater Surveillance System
CFSAN: Center for Food Safety and Applied Nutrition
C-WAP: CFSAN Wastewater Analysis Pipeline
ONT: Oxford Nanopore Technologies
NFW: nuclease-free water
RNA: ribonucleic acid
SNV: single nucleotide variant
NCBI: National Center for Biotechnology Information
PCR: polymerase chain reaction
ddPCR: droplet digital PCR
Pangolin: Phylogenetic Assignment of Named Global Outbreak Lineages
VOC: variant of concern
DCIPHER: Data Collation and Integration for Public Health Event Responses
S3C: Swiss SARS-CoV-2 Sequencing Consortium
SIB: Swiss Institute of Bioinformatics

