## Supplemental Materials for "A Gold Standard Dataset for Lineage Abundance Estimation from Wastewater"

Corresponding Author :

**Supplemental Table 1: Composition of Synthetic RNA control mixtures**

| Mixture | Controls | Concentration (Copies/ul) |
|---|---|---|
| All | Wuhan hu-1 from China, Wuhan hu-1 from California, Alpha, Gamma, Iota, Delta, Omicron - BA.1 lineage, B.1.1.529+BA.2-England, B.1.1.529+BA.2-Australia, B.2.12.1-Denmark, B.2.12.1-USA, BA.5-England, BA.5-USA, BA.4-Texas, BA.4-California, NFW | 625 |
| ØADGIO1 | Wuhan hu-1 from china,<br>Wuhan hu-1 from California,<br>Alpha,<br>Gamma,<br>Iota,<br>Omicron - BA.1 lineage,<br>Delta | 1875<br>625<br>2500<br>625<br>1250<br>1875<br>1250 |
| O2O3O4O5 | B.1.1.529+BA.2-Australia,<br>B.1.1.529+BA.2-England,<br>B.2.12.1-Denmark,<br>B.2.12.1-USA,<br>BA.5-England,<br>BA.5-USA,<br>BA.4-Texas,<br>BA.4-California | 1250<br>625<br>1875<br>625<br>2500<br>625<br>1250<br>1250 |
| ØAGIO1O2 | Wuhan hu-1 from china,<br>Wuhan hu-1 fromCalifornia,<br>Alpha,<br>Gamma,<br>Iota,<br>B.1.1.529+BA.2-Australia,<br>Omicron - BA.1 lineage,<br>B.1.1.529+BA.2-England | 625<br>625<br>625<br>1875<br>1250<br>1875<br>1875<br>1250 |

| | | |
|---|---|---|
| ØO5O3O4 | Wuhan hu-1 from China,<br>Wuhan hu-1 from California,<br>B.2.12.1-Denmark,<br>B.2.12.1-USA,<br>BA.5-England,<br>BA.5-USA,<br>BA.4-Texas,<br>BA.4-California | 2500<br>1250<br>1250<br>625<br>1250<br>1250<br>625<br>1250 |
| ADGIO1O2O3 | Alpha,<br>Gamma,<br>Iota,<br>Delta,<br>Omicron - BA.1 lineage,<br>B.1.1.529+BA.2-England,<br>B.1.1.529+BA.2-Australia,<br>B.2.12.1-Denmark | 625<br>625<br>625<br>625<br>1875<br>1875<br>1250<br>2500 |
| AGIO3O4O5 | Alpha,<br>Gamma,<br>Iota,<br>B.2.12.1-USA,<br>BA.5-England,<br>BA.5-USA,<br>BA.4-Texas,<br>BA.4-California | 1250<br>1250<br>1875<br>1250<br>1875<br>625<br>625<br>1250 |
| O1O2O3O4O5 | Omicron - BA.1 lineage,<br>B.1.1.529+BA.2-Australia,<br>B.1.1.529+BA.2-England,<br>B.2.12.1-Denmark,<br>BA.5-USA,<br>BA.4-California | 2500<br>1250<br>1250<br>1250<br>2500<br>1250 |
| Ø | Wuhan hu-1 from China,<br>Wuhan hu-1 from California | 3750<br>6250 |
| O1O2 | Omicron - BA.1 lineage,<br>B.1.1.529+BA.2-Australia,<br>B.1.1.529+BA.2-England | 3750<br>5000<br>1250 |
| O3 | B.2.12.1-Denmark, B.2.12.1-USA | 5000 |
| O5 | BA.5-England,<br>BA.5-USA | 7500<br>2500 |
| O4 | BA.4-Texas, | 6250<br>3750 |

| | | |
|---|---|---|
| | BA.4-California | |
| Ø-2 | Wuhan hu-1 from California | 10000 |
| A | Control Alpha | 10000 |
| G | Control Gamma | 10000 |
| I | Iota | 10000 |
| D | Control Delta | 10000 |
| Ø-3 | Wuhan hu-1 from China | 10000 |
| O3-2 | B.2.12.1-Denmark | 10000 |
| O3-3 | B.2.12.1(USA) | 10000 |
| O5-2 | BA.5-England | 10000 |
| O5-3 | BA.5-USA | 10000 |
| O4-2 | BA.4-Texas | 10000 |
| O4-3 | BA.4-California | 10000 |
| O2-2 | B.1.1.529+BA.2-Australia | 10000 |
| O2O3O4O5-2 | B.1.1.529+BA.2-England, B.2.12.1(USA), BA.5-USA, BA.4-California | 2500 |
| O2O3O4O5-3 | B.1.1.529+BA.2-Australia, B.2.12.1-Denmark, BA.5-England, BA.4-Texas | 2500 |
| ØADGIO1-2 | Wuhan hu-1 from China, Wuhan hu-1 from California, Alpha, Gamma, Iota, Delta, Omicron - BA.1 lineage | 1425 |
| ØAIO1O2O3O4O5 | Wuhan hu-1 from China, Alpha, Iota, Omicron - BA.1 lineage, B.1.1.529+BA.2-Australia, B.2.12.1(USA), BA.5-England, BA.4-California | 1250 |
| Ø-4 | Wuhan hu-1 from China, NFW | 7500 |

| | | |
|---|---|---|
| A-2 | Control Alpha, NFW | 7500 |
| G-2 | Control Gamma, NFW | 7500 |
| I-2 | Iota, NFW | 7500 |
| D-2 | Control Delta, NFW | 7500 |
| O1-2 | Control Omicron - BA.1 lineage, NFW | 7500 |
| O2-3 | B.1.1.529+BA.2-England, NFW | 7500 |
| O3-4 | B.2.12.1(USA), NFW | 7500 |

**Supplemental Materials 1.**

**Tool usage and explanation of deviation from default parameters:**

In an attempt to make all of these deconvolution tools comparable, a few tweaks to configuration files or input parameters were made. Below, we have detailed any deviations from the normal usage described on each tool's GitHub page. A repository dedicated to showing precisely how we implemented each tool can be found at - https://github.com/enviro-lab/benchmark-deconvolute.

**Alcov:** https://github.com/Ellmen/alcov

Default settings were used for Alcov to determine relative abundance. For input, it requires a folder of bam files and a .txt file that has a list of the paths for each of the .bam files and their names.

**Freyja:** https://github.com/andersen-lab/Freyja

Our Freyja results came from running the CDC's C-WAP pipeline, which uses default settings for the `freyja variants` step and employs the --confirmedonly flag in the `freyja demix` step in order to exclude unconfirmed lineages.

**Kallisto:** https://github.com/pachterlab/kallisto

As a repurposed metagenomics tool, kallisto requires the user to create a new index for use in lineage determination. A randomized selection of 100 lineages was chosen for each lineage with at least 500 occurrences in a USA subset of GISAID metadata (Khare et al, 2021) downloaded in

September of 2023, and the associated fasta sequences were fed into the pipeline described in the GitHub repo wastewater_analysis (Baaijens, 2021) which uses `kallisto index` to create an index file from the reference set before quantifying lineage proportions present in each sample.

**Lineagespot:** https://github.com/npechl/lineagespot

While lineagespot includes reference files for 13 pangolin lineages, we needed to be able to predict the presence of lineages not among that group. We added in over 2000 other lineages that had appeared at high prevalence levels at any time in the USA, as reported by in the GISAID metadata described in the kallisto section above. Lineages were downloaded using R-outbreak-info's (Outbreak-Info, 2023) tool getMutationsByLineage and were converted to Lineagespot's input format using a custom script. Lineagespot relies on allele frequency details from the input .vcf file. ARTIC .bam files were converted to .vcf files with bcftools and SnpEff (Cingolani, 2012) was used to annotate the .vcf to the required format. The output was filtered so that only mutations present in the original ARTIC .vcf remained in the final .vcf provided to Lineagespot.

**LCS:** https://github.com/rvalieris/LCS

To download recent, public mutation information, LCS was run with the config setting markers=ucsc, and the file LCS/rules/config.py was edited so that PB_VERSION='2023-07-24' and PRIMERS_FA='data/V4_primers.fa', where the primers fasta contained sequences corresponding to the ARTIC v4 primers used in sequencing; this provided twenty-six lineages. LCS outputs lineages with the WHO lineage name (if available) separated by an underscore from the pangolin lineage assignment (e.g. Gamma_P.1 or AV.1) and occasionally trailed with another sublineage (e.g. Epsilon_B.1.427_429). Lineages like the latter example where the pangolin lineage was unclear were considered in part of the "Other" category, which contained any unassigned lineages, but otherwise, the pangolin lineage was used for analyses.

**LolliPop:** https://github.com/cbg-ethz/lollipop

LolliPop differs from most of the other tools in that it considers time series. Given that some of our mixtures combine lineages that did not exist at the same time as each other, LolliPop was configured to ignore time by adding "no_date: True" to the variants-config file. Because LolliPop is interested in portraying lineage proportions for a given location, each sample name was used as a different location in the locations_list of the variants-config file, and location and sample name were used interchangeably in further analyses. LolliPop outputs lower and upper bounds along with its estimated lineage proportions; only the proportions were considered when aggregating results to compare with other tools. Like Lineagespot, LolliPop's initial reference set is small, only containing eleven lineages, so additional lineages were acquired as recommended by the LolliPop authors from PHE Genomic's Standardised Variant Definitions (SVDs) (Bull, et al, 2023), which lists thirty-four lineages (including the original eleven), and `cojac phe2cojac` (Jahn et al, 2022) was used to convert to the format used by LolliPop. Because of the small size of the SVDs, LolliPop had the least comprehensive dataset of lineages to detect compared to the other tools.

**VaQuERo:** https://github.com/fabou-uobaf/VaQuERo

Like LolliPop, VaQuERo, by default, considers location and date in lineage assignment. To avoid this behavior, the parameter --smoothingsamples was set to 0, and the dates for each mixture in the metadata provided to VaQuERo were all set three days apart, since all locations were set equivalent. VaQuERo relies on allele frequency details from the input .vcf file. Since the ARTIC pipeline does not include this in the output .vcf, each mixture's .bam output from ARTIC was run through LoFreq (Wilm et al, 2012), a variant caller mentioned in the VaQuERo documentation, which outputs allele frequencies in the output .vcf, and this file was used for analysis.

**Kallisto and Kraken 2 (C-WAP):** https://github.com/CFSAN-Biostatistics/C-WAP

As part of C-WAP, kallisto and a combination of Kraken 2 and Bracken were separately employed as alternative deconvolution tools. Both had a database created using a few consensus sequences for each of several lineages listed by Kayikcioglu, et al., 2023. These tools were used unaltered from their implementation in C-WAP.

**Standardization of outputs**

Each tool compared in this analysis had its own output format. To make our outputs comparable, they have been converted to the same output format used by Freyja. Pangolin lineages provided by each tool were summarized in the same manner derived from the way Freyja summarized lineages, but with a few categories adjusted to best illustrate the proportions of relevant lineages and sublineages; most notably, BA.1, BA.2, BA.4, and BA.5 and their sublineages were grouped separately rather than remaining within a single, broad Omicron category.

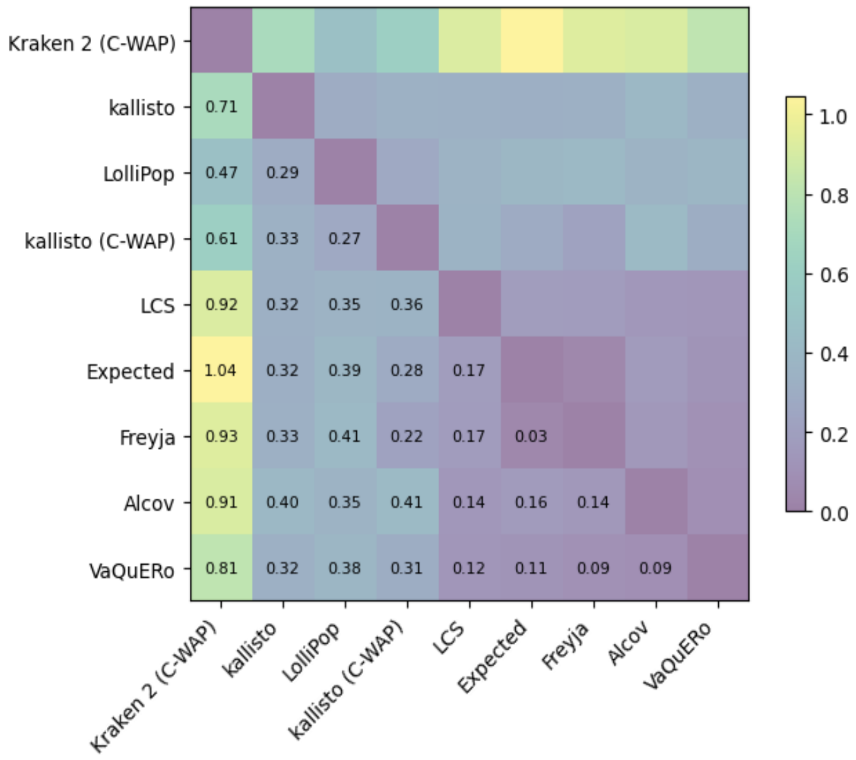


**Supplemental Figure 1:** Median pairwise L2 abundance norms between deconvolution tools (Varskip Dataset)

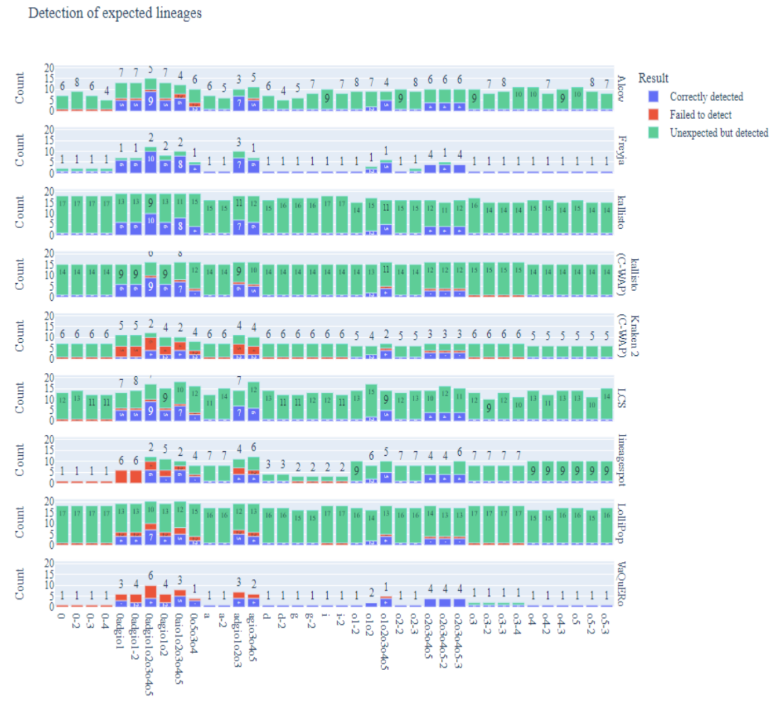

**Supplemental Figure 2:** Detection of expected lineage by the deconvolution tools (WB mixtures-Varskip dataset)

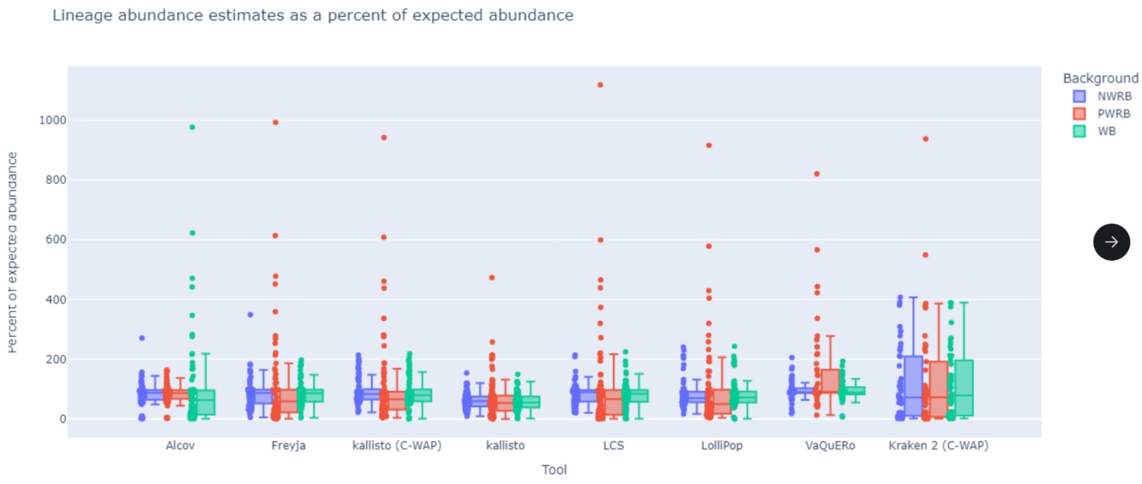


**Supplemental Figure 3:** Comparison of relative abundance of datasets in different backgrounds (Varskip Dataset)

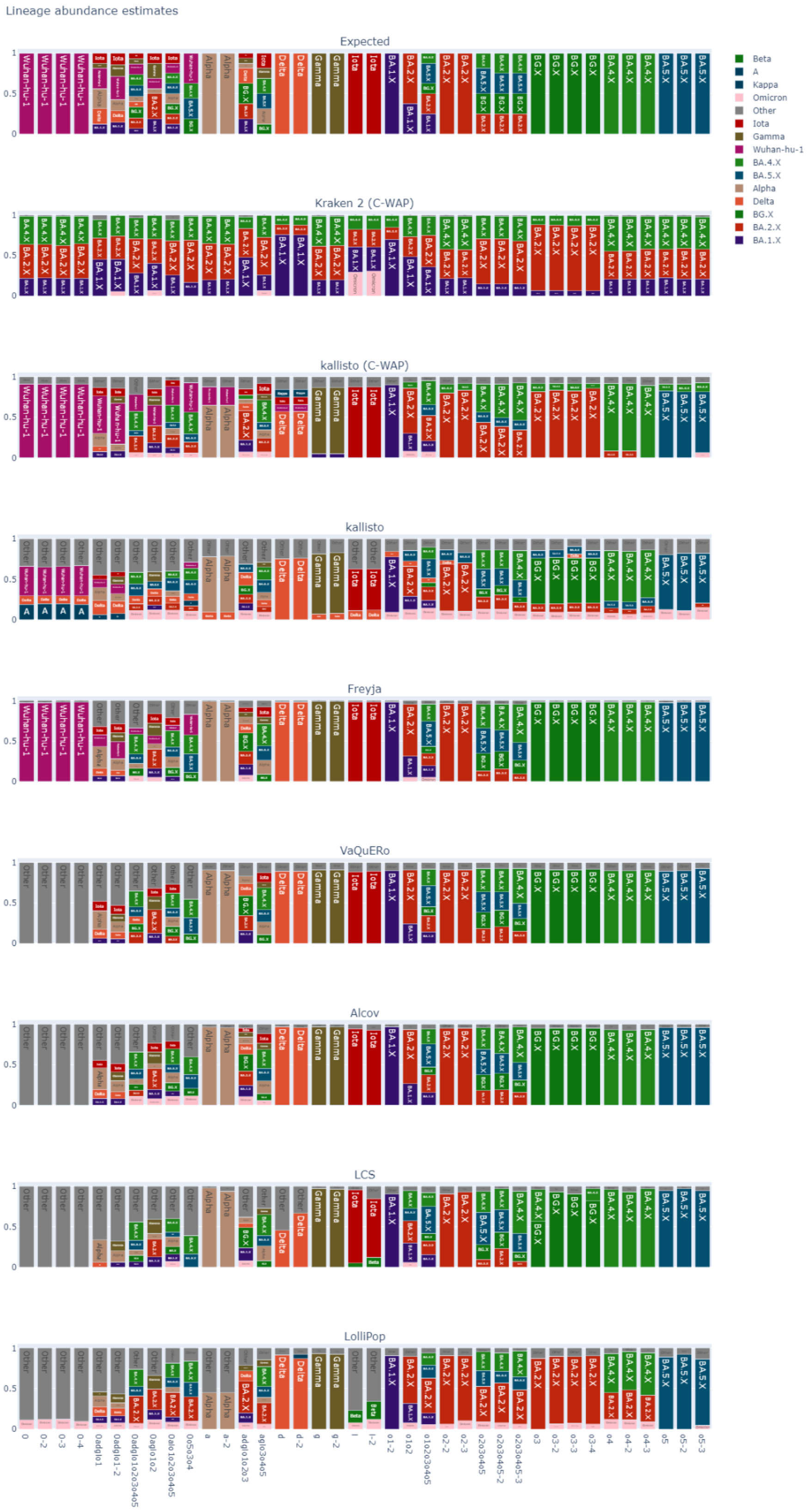


**Supplemental Figure 4:** Lineage abundance estimation of different tools with Artic dataset

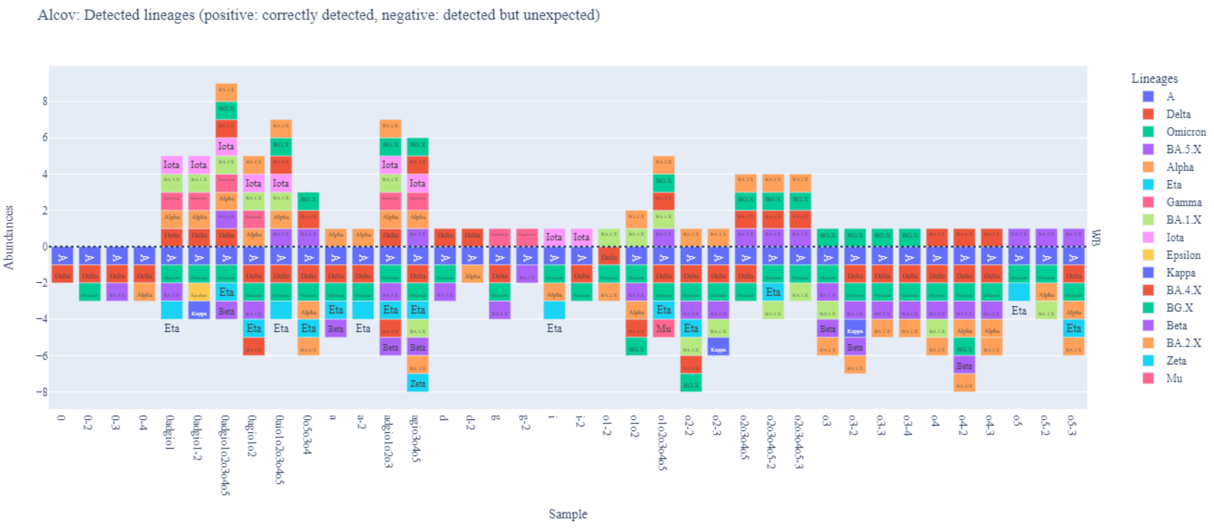
Alcov: Detected lineages (positive: correctly detected, negative: detected but unexpected)
Abundances
Sample
Lineages
A
Delta
Omicron
BA.5.X
Alpha
Eta
Gamma
BA.1.X
Iota
Epsilon
Kappa
BA.4.X
BG.X
Beta
BA.2.X
Zeta
Mu

5a

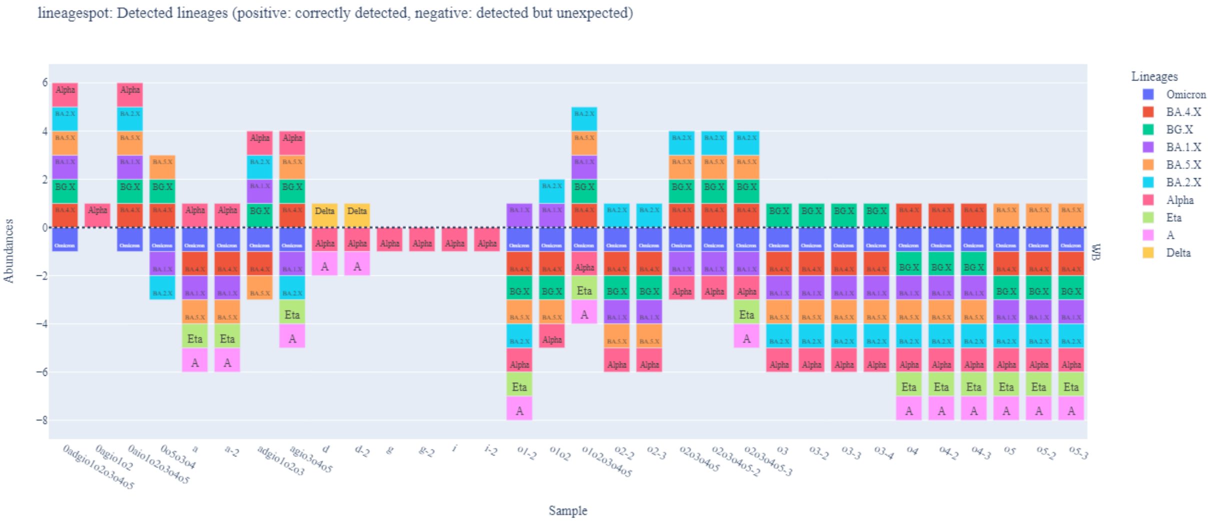
lineagespot: Detected lineages (positive: correctly detected, negative: detected but unexpected)
Abundances
Sample
Lineages
Omicron
BA.4.X
BG.X
BA.1.X
BA.5.X
BA.2.X
Alpha
Eta
A
Delta

5b

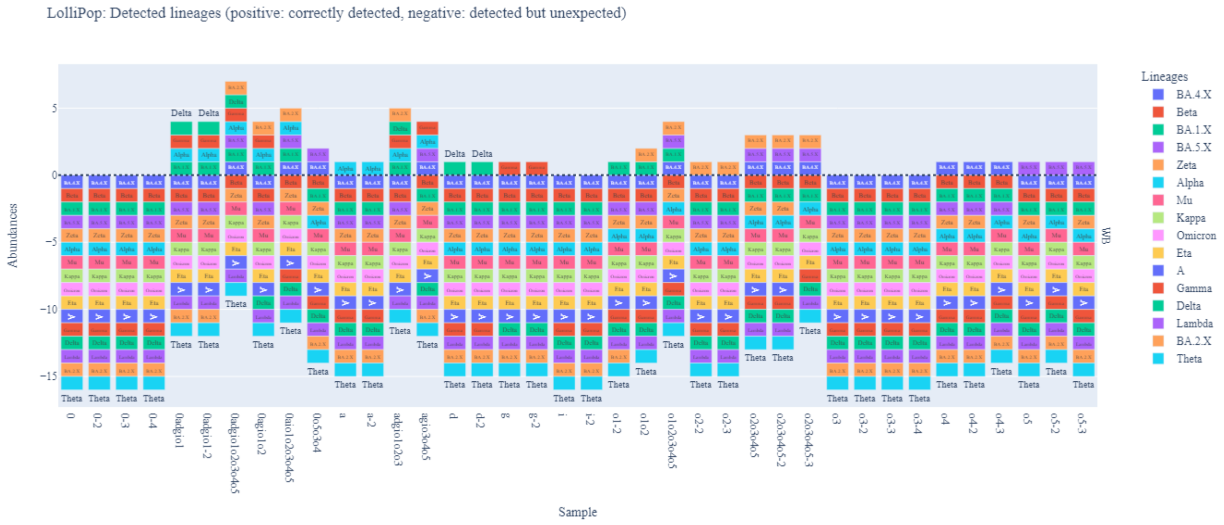
LolliPop: Detected lineages (positive: correctly detected, negative: detected but unexpected)
Abundances
Sample
Lineages
BA.4.X
Beta
BA.1.X
BA.5.X
Zeta
Alpha
Mu
Kappa
Omicron
Eta
A
Gamma
Delta
Lambda
BA.2.X
Theta
WB

5c

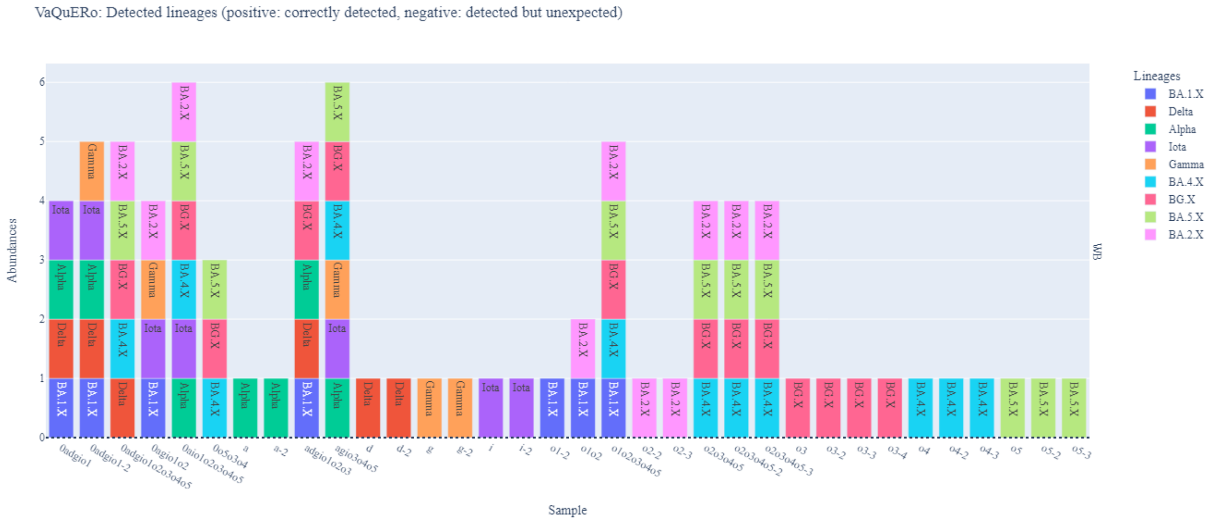
VaQuERo: Detected lineages (positive: correctly detected, negative: detected but unexpected)
Abundances
Sample
Lineages
BA.1.X
Delta
Alpha
Iota
Gamma
BA.4.X
BG.X
BA.5.X
BA.2.X
WB

5d

kallisto (C-WAP): Detected lineages (positive: correctly detected, negative: detected but unexpected)

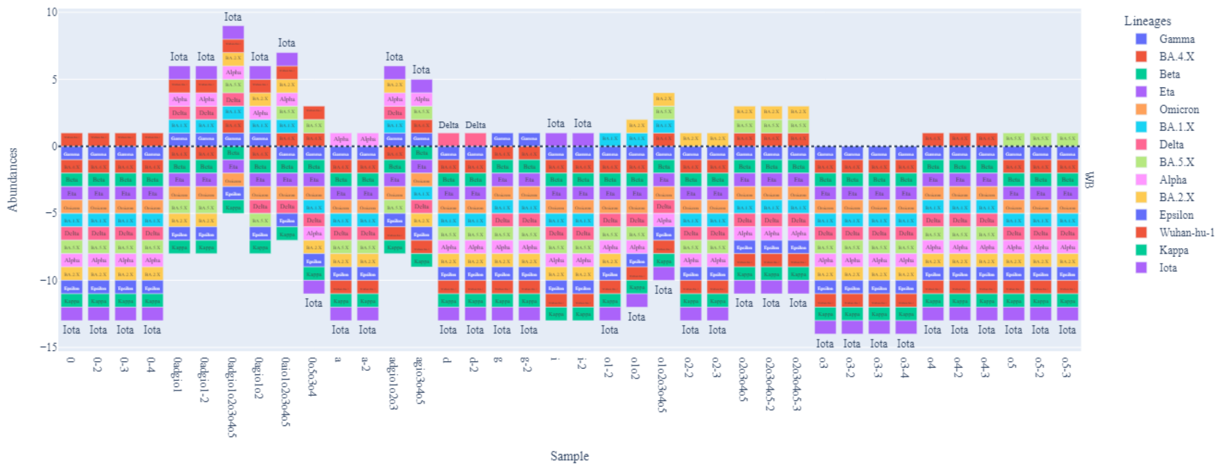


kallisto: Detected lineages (positive: correctly detected, negative: detected but unexpected)

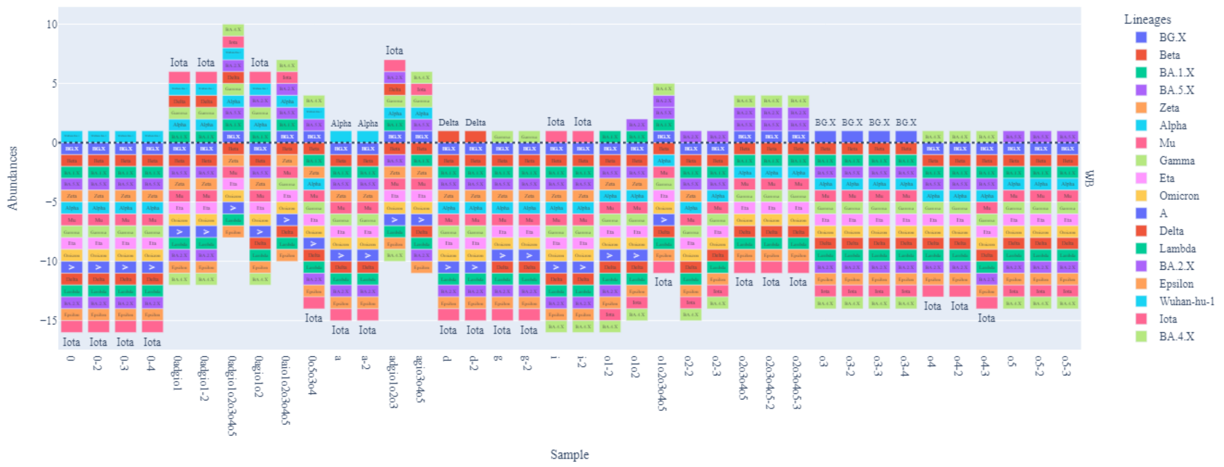


5e

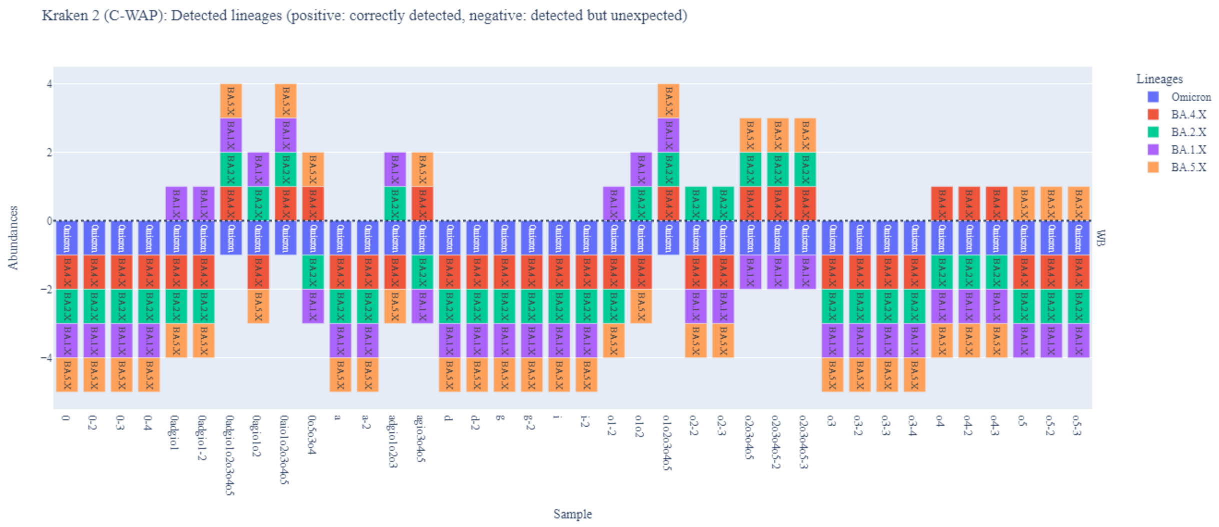


5f

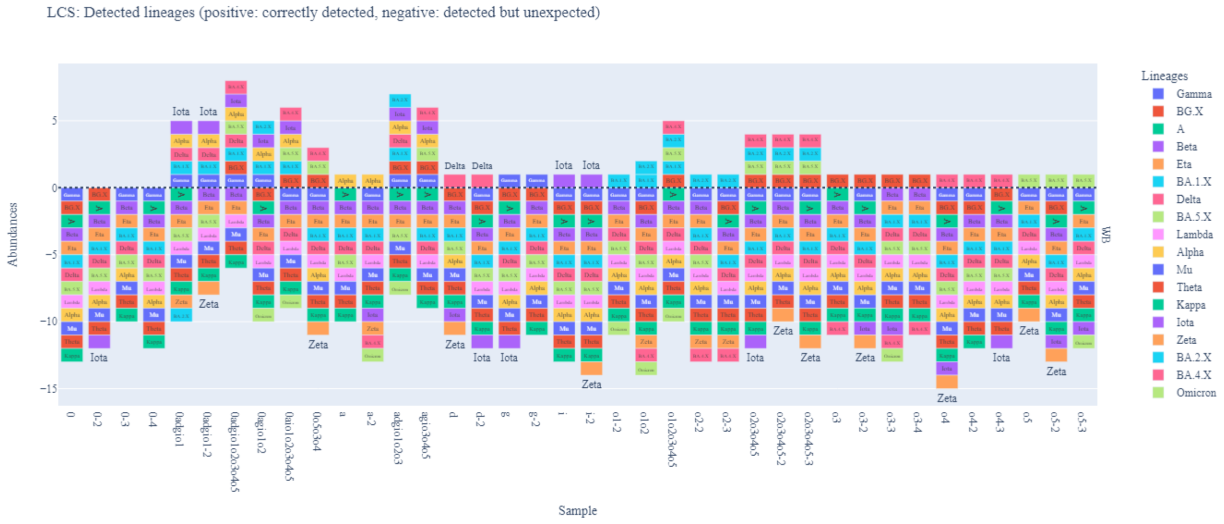


5g

**Supplemental Figure 5:** Detected lineage composition by different deconvolution tools (WB mixtures) - a)Alcov, b)lineageSpot, c)LolliPop, d)VaQuERo, e)Kallisto with two different datasets, f)Kraken, g)LCS. Results are ordered from left to right by the first date of appearance of the VOCs included in sample mixtures prepared for this study. Samples containing the original Wuhan strain of SARS-CoV-2 thus appear on the far left and samples containing Omicron sub lineages appear on the far right.
